# PET/CT guided tuberculosis treatment shortening: a randomized trial

**DOI:** 10.1101/2024.10.03.24314723

**Authors:** Stephanus T. Malherbe, Ray Y. Chen, Xiang Yu, Bronwyn Smith, Xin Liu, Jingcai Gao, Andreas H. Diacon, Rodney Dawson, Michele Tameris, Hong Zhu, Yahong Qu, Hongjian Jin, Shouguo Pan, Lori E. Dodd, Jing Wang, Lisa C. Goldfeder, Ying Cai, Kriti Arora, Joel Vincent, Kim Narunsky, Keboile Serole, Rene T. Goliath, Laylah Da Costa, Arshad Taliep, Saalikha Aziz, Remy Daroowala, Friedrich Thienemann, Sandra Mukasa, Richard Court, Bianca Sossen, Petri Ahlers, Simon C. Mendelsohn, Lisa White, Aurélie Gouel, Chuen-Yen Lau, Samy Hassan, Lili Liang, Hongfei Duan, Gita K. Moghaddam, Praveen Paripati, Saher Lahouar, Michael Harris, Kurt Wollenberg, Brendan Jeffrey, Mike Tartakovsky, Alex Rosenthal, Michael Duvenhage, Derek T. Armstrong, Taeksun Song, Jill Winter, Qian Gao, Laura E. Via, Robert J. Wilkinson, Gerhard Walzl, Clifton E. Barry

**Author notes:** These authors contributed equally to this work (Stephanus T. Malherbe, Ray Y. Chen). Corresponding authors. (G.W.); (C.E.B.).

## Abstract

**Six months of chemotherapy using current agents is standard of care for pulmonary, drug-sensitive tuberculosis (TB), even though some are believed to be cured more rapidly and others require longer therapy. Understanding what factors determine the length of treatment required for durable cure in individual patients would allow individualization of treatment durations, provide better clinical tools to determine the of appropriate duration of new regimens, as well as reduce the cost of large Phase III studies to determine the optimal combinations to use in TB control programs. We conducted a randomized clinical trial in South Africa and China that recruited 704 participants with newly diagnosed, drug-sensitive pulmonary tuberculosis and stratified them based on radiographic disease characteristics as assessed by FDG PET/CT scan readers. Participants with less extensive disease (N=273) were randomly assigned to complete therapy after four months or continue receiving treatment for six months. Amongst participants who received four months of therapy, 17 of 141 (12.1%) experienced unfavorable outcomes compared to only 2 of 132 (1.5%) who completed six months of treatment (treatment success 98.4% in B, 86.7% in C (difference -11.7%, 95% CI, -18.2%, -5.3%)). In the non-randomized arm that included participants with more extensive disease, only 8 of 248 (3.2%) experienced unfavorable outcomes. Total cavity volume and total lesion glycolysis at week 16 were significantly associated with risk of unfavorable outcome in the randomized participants. Based on PET/CT scans at TB recurrence, bacteriological relapses (confirmed by whole genome sequencing) predominantly occurred in the same active cavities originally present at baseline. Automated segmentation of the serial PET/CT scans was later performed, and machine-learning was used to classify participants according to their likelihood of relapse, allowing the development of predictive models with good performance based on CT, PET, microbiological and clinical characteristics. These results open the possibility for more efficient studies of future TB treatment regimens.**

## Introduction

Treatment of drug-susceptible tuberculosis (TB) with the agents currently in use requires at least six months of treatment to achieve durable cure (defined as failure to relapse after drug discontinuation)(*1, 2*). Patients receive an initial two month “intensive phase” with four drugs daily (isoniazid, rifampicin, pyrazinamide and ethambutol) followed by 4 months of therapy with two of these agents (isoniazid and rifampicin). Failure to convert sputum smear or culture by two months or the presence of cavities on chest X-ray at baseline are used in high resource countries as a basis for extending the two drug “continuation phase” by an additional 3 months to 9 full months. Many high TB burden countries follow the guidelines of the World Health Organization which stipulate 4-6 months of daily therapy for all TB patients(*3*).

The pioneering trials of the British Medical Research Council that led to the establishment of this standard of care regimen involved exploration of the duration of both the intensive and continuation phases and noted a curious phenomenon that about 80% of patients could be treated for only three months and be cured of their disease but to cure the remaining 20% the duration of therapy had to be extended for a full six months(*4*). This bimodal distribution of treatment responses suggests an underlying difference in the disease characteristics of patients cured early in treatment compared to those who require extended treatment times. One characteristic that could be different between these groups is the presence and extent of cavitary disease which has been associated with disease relapse in many studies(*5-9*). Both the presence of cavities and cavity size correlate with higher sputum bacterial load and disease severity(*10-13*). Another characteristic that clearly associated with risk of relapse is the susceptibility of individual strains to low concentrations of rifampicin and isoniazid, concentrations that are not high enough to meet clinically established breakpoints to designate them as resistant but nonetheless potentially conferring survival benefit in some niches in patients(*14*).

Until the success of the TBTC Study 31 trial(*15*), no trial testing a 4-month treatment arm achieved non-inferiority compared to the 6-month standard of care; this included the older BMRC trials as well as, the more recent fluoroquinolone-based trials(*16-18*). These trials enrolled participants without regard to disease severity, with the three fluoroquinolone-based studies having unfavorable outcomes ranging from 15-20% relapse in the 4-month arms. In contrast, Johnson et al conducted a 4-month treatment shortening trial in which only participants with less severe disease, defined as no cavity on baseline chest x-ray and negative week 8 sputum culture, were randomized to 4 vs 6 months of treatment(*19*). This study revealed a 7.0% unfavorable outcome rate in the 4-month arm, driven largely by African participants, which was significantly worse than the 1.6% unfavorable outcome rate in the 6-month arm.

To identify the biological basis for the elevated risk of relapse disease in some patients, we conducted a prospective, randomized controlled noninferiority trial of four versus six months of chemotherapy in pulmonary TB patients with a combination of radiographic characteristics at baseline, the amount of change of these features at one month, and markers of residual bacterial load at the end of treatment.

## RESULTS

### Participant flow and randomization

We screened 972 participants, enrolled a total of 704 of whom 558 were available for stratification or randomization (Figure 1). Based on high baseline cavity and lesion burden, deterioration from baseline to week 4 (determined by repeat PET/CT), or week 16 sputum GeneXpert MTB/RIF cycle threshold values ≥28, 250 participants (44%) were not eligible for randomization and were stratified to receive standard treatment for 24 weeks (Arm A). Two of these participants were subsequently found to have drug-resistance and were withdrawn. 276 (49%) participants were eligible for randomization to receive either 24 weeks of treatment (Arm B, 135 participants, 3 of whom were subsequently found to have baseline drug-resistance and withdrawn), or to stop treatment after 16 weeks (Arm C, 141 participants). During interim analysis, the Data Safety and Monitoring Board reviewed the results and stopped recruitment into the 16-week treatment arm due to meeting pre-specified stopping criteria for inferiority compared to 24-weeks of treatment. Thirty-two participants who were enrolled in the study at this time but not yet randomized, and were not stratified to Arm A, were subsequently assigned to Arm B.

**Figure 1.**
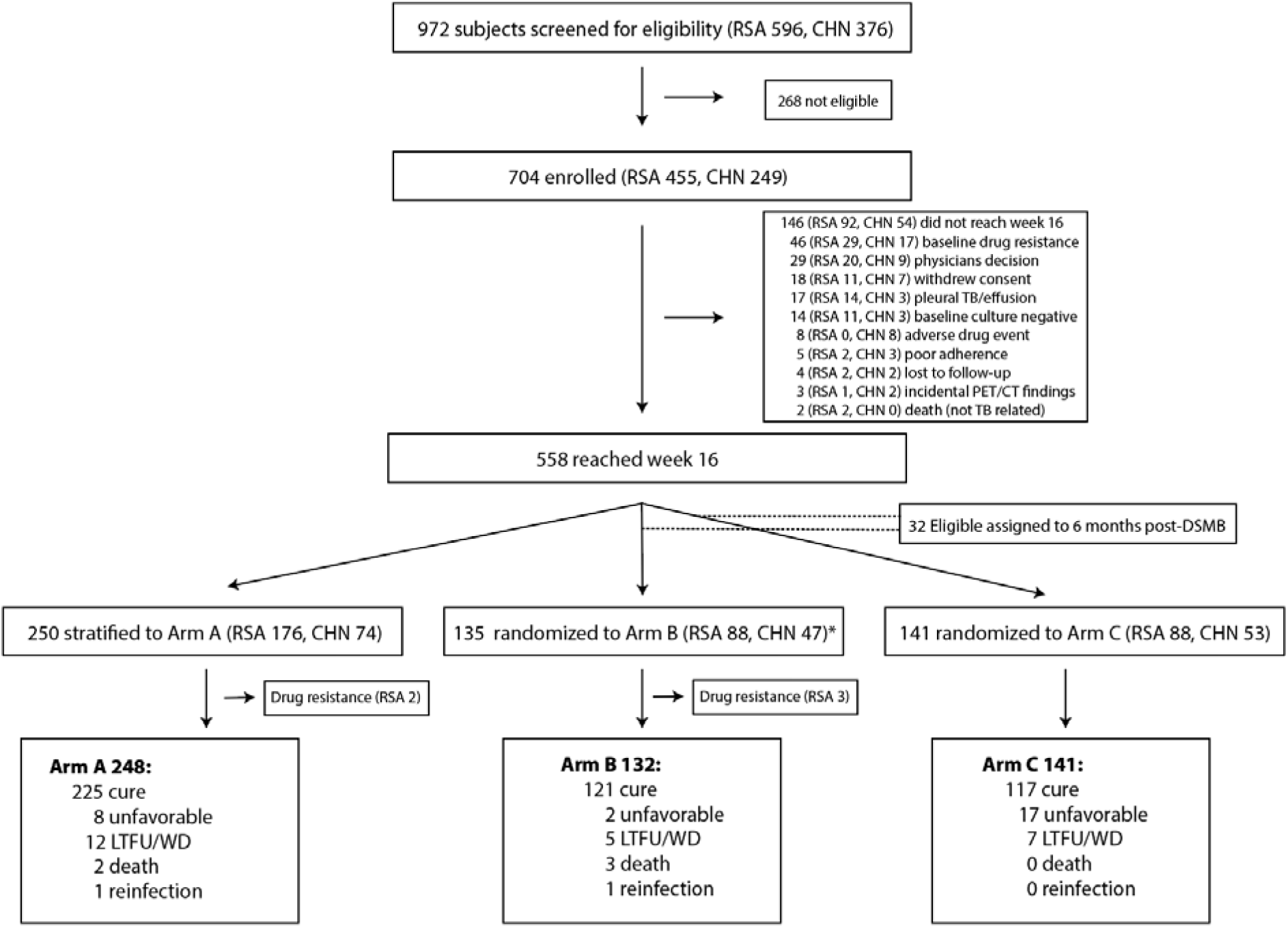
CONSORT recruitment flow diagram. Of 972 participants screened, 704 were enrolled and 146 were withdrawn from the study for the reasons shown (Details of reasons for ineligibility are given in Supplemental Table 1). Of these, 248 were stratified to the non-randomized arm (A) due to the disease severity and 276 were randomly assigned to Arm B or Arm C. *After the DSMB stopped randomization an additional 32 (11 RSA, 21 CHN) participants in the Arm B/C pool were assigned to Arm B (not shown in figure).

Baseline characteristics did not differ significantly by study arm except for radiologic features (cavities, hard CT volume, and total lesion glycolysis (TLG)). Participants assigned to Arm A, by design, had larger and more numerous cavities, larger CT hard volumes, TLG and higher bacterial burden (Table 1). By country, baseline characteristics were significantly different in that Chinese participants had significantly less extensive disease with fewer and smaller cavities, lower TLG, and higher bacterial burdens. Consistent with this, amongst participants assigned to Arm A, 70.2% were from South Africa and only 29.8% were Chinese. Participants were assigned to Arm A primarily because of baseline TLG exceeding 1500 followed by baseline hard volume greater than 200 mL and week 16 GeneXpert Ct values less than 28 (Supplemental Figure 1).

**Table 1.**
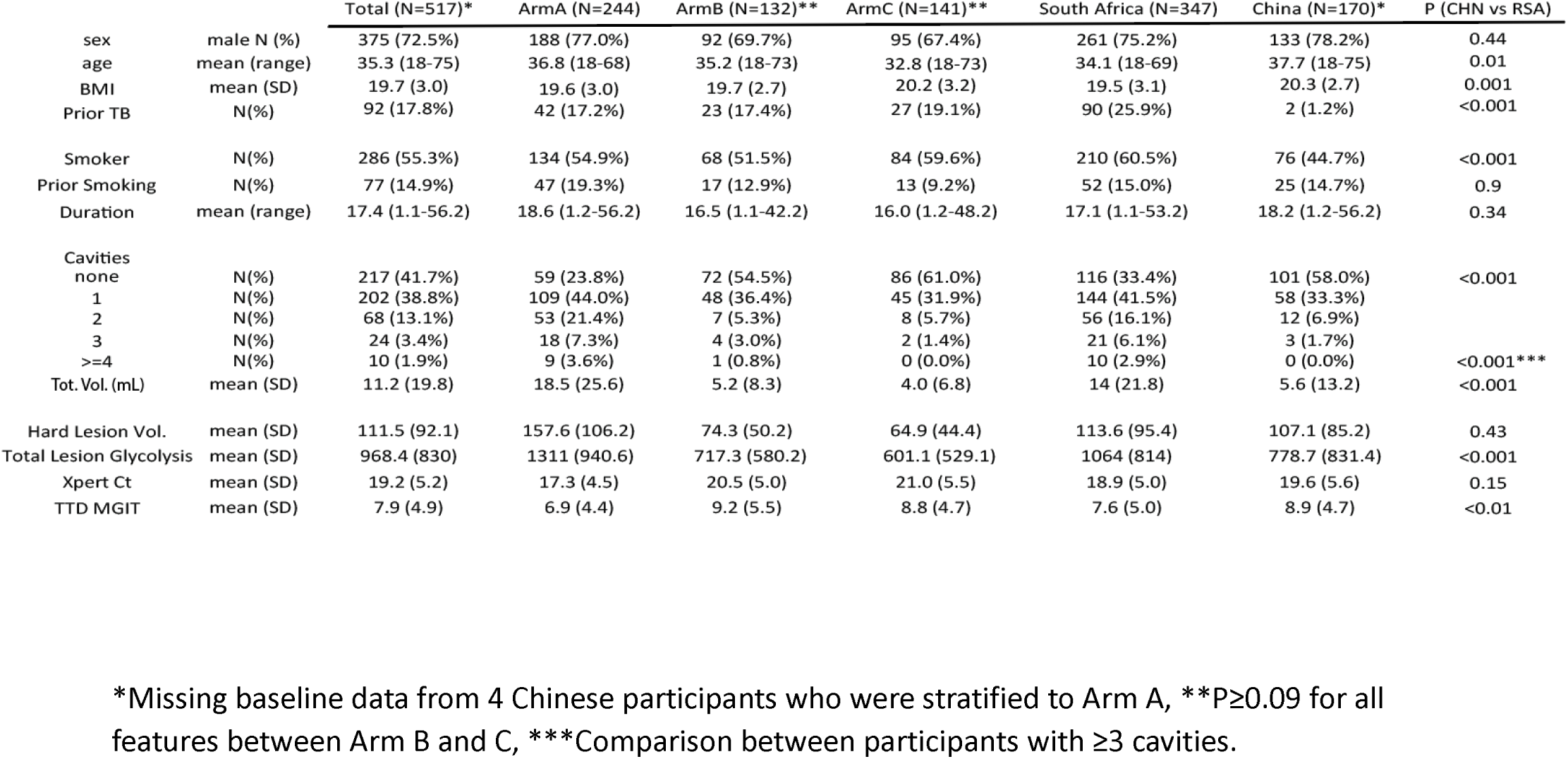
Baseline subject characteristics by Arm and county. BMI: Body Mass Index, Xpert Ct: GeneXpert cycle threshold, TTD MGIT: Time to Detection in the MGIT (Days). Cavities were defined as airspace >2mL within the lung parenchyma with an interior HU value of <-750HU on CT scan.

### Final outcomes

Overall treatment outcomes at the end of the study were assessed as shown in Fig. 2B. These results showed that participants in Arm C had significantly more unfavorable outcomes (confirmed failure and relapse cases) than participants in Arm B (primary endpoint) or Arm A (secondary endpoint; Figure 2A). Notably participants in Arm A, deemed to have more severe disease, following 6 months of treatment only experienced 7 relapses and one treatment failure out of 248 participants (3.2%). In Arm B, which included 132 participants with less severe disease who were randomized to the full 6 months of treatment, there were only 2 relapses (1.5%) and no failed treatment cases. This was only slightly lower than Arm A. In the four-month treatment arm, Arm C, amongst 141 participants there were 14 relapses and 3 treatment failures (12.1%).

**Figure 2.**
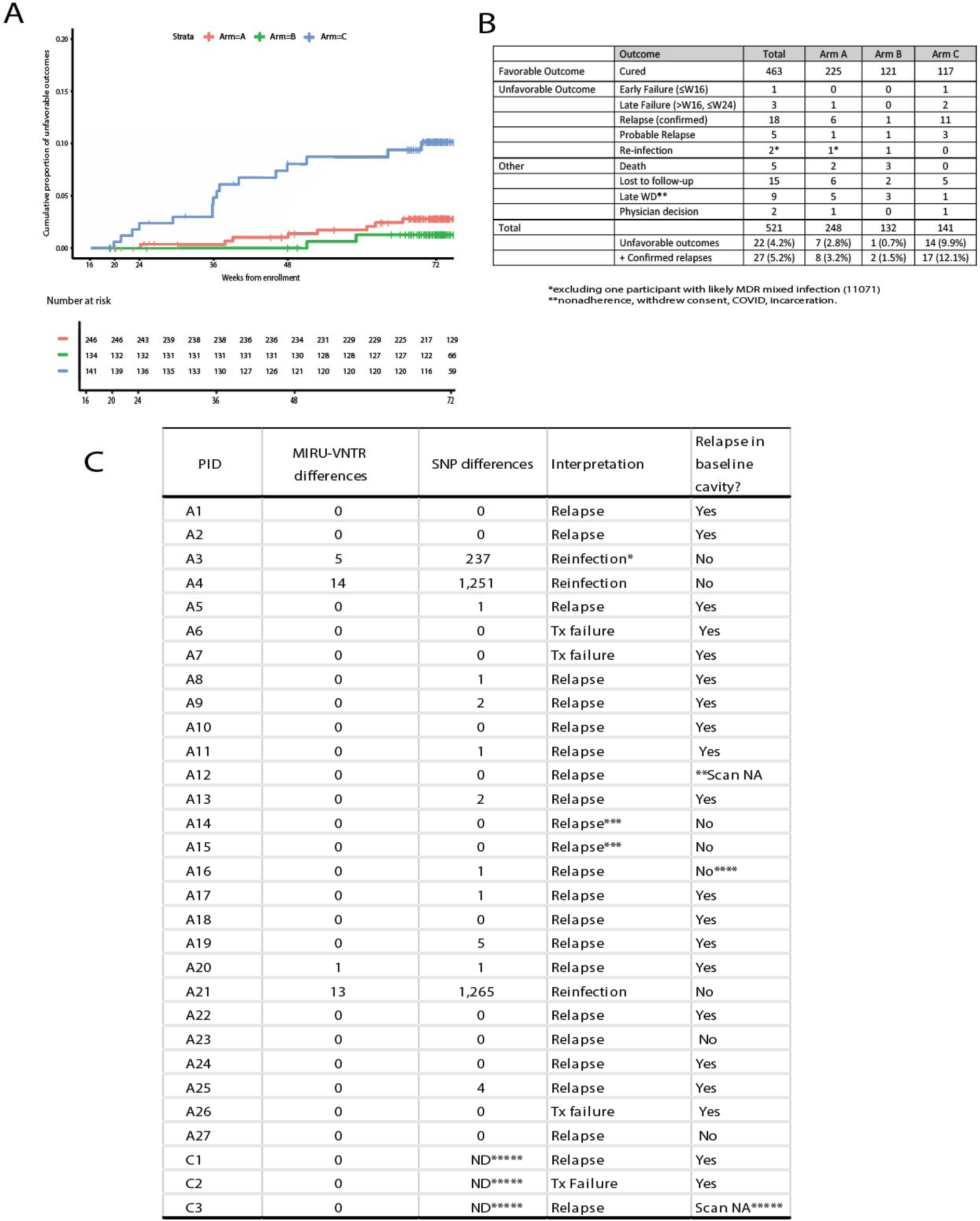
Final study outcomes. **A.** Kaplan-Meier plot showing the cumulative proportion of unfavorable outcomes starting at week 16, the time of randomization. **B.** Table showing participant final outcomes by Arm. **C.** Table of unfavorable outcomes with genomic information, interpretation and whether the relapse involved a cavity present at baseline. *The week 16 isolate was MDR, which may have been from a mixed infection initially, **Subject had a stroke and was not able to be scanned, ***A14 and A15 were relatives and cohabitated during the study, re-infection with the same strain cannot be definitively ruled out (Supplemental Figure 2), ****relapse in lower lobe adjacent to cavity that infiltrated the fissure, *****WGS data and final scan not available for these Chinese participants…

Recurrent disease was assessed by MIRU-VNTR genotyping and whole genome sequencing of the bacterial isolate from baseline compared to a sputum culture isolate obtained at the time of recurrence (Figure 2C). Amongst the 30 failed treatment and recurrence cases, 27 were observed from South Africa. Three failed treatment cases showed 0 SNP differences when comparing the baseline and final isolate, 21 relapses contained between 0-5 SNP differences when comparing baseline and recurrent isolates, and 3 participants were determined to have been reinfected based on between 237-1,265 SNP differences between their baseline and recurrent isolates (Figure 2C). For the three unfavorable outcomes from China only MIRU-VNTR genotyping data were available but both relapses and one treatment failure showed no change in MIRU-VNTR pattern. None of the observed SNPs in relapse strains was linked with acquisition of drug-resistance.

### Primary and secondary endpoint analysis

For the primary endpoint treatment success was 98.4% in B and 86.7% in C (difference -11.7%, 95% CI, -18.2%, -5.3%). We compared clinical, microbiological and radiological characteristics between patients in the two randomized Arms B and C to determine risk factors that correlated with poor outcomes. In univariate Cox proportional hazards analysis, assignment to the four-month early discontinuation arm (C) was the strongest predictor of unfavorable outcome with a HR of 7.99 (95% confidence interval (CI) 1.84 to 34.7) (Table 2). Many features associated with cavities also showed significant p-values in this analysis including cavity volume at week 16 and percent change in cavity volume from week 0 to week 16. Total lesion glycolysis at week 4 and 16, change of total lesion glycolysis from week 0 to 16(log ratio), GeneXpert cycle threshold at baseline and week 16 were also significantly associated with unfavorable outcome. Surprisingly however, hard lesion volume (HU -100 to +200) did not correlate well with outcome, either as absolute values or as changes from baseline. In the multivariate analysis, assignment to Arm C remained the dominant risk factor but total cavitary volume at week 16 and total lesion glycolysis at week 16 also remained significant. Adding participants from Arm A (who were stratified to all receive six months of treatment) into the analysis strengthened the association between 4-month treatment and unfavorable outcome, with total lesion glycolysis at week 16 and BMI at baseline also contributing significantly to the multivariate model. Finally, to remove the large effect of treatment duration we performed an analysis of only those participants randomized to Arm C. In the multivariate model, total lesion glycolysis at week 16 and BMI at baseline remained the primary correlates of unfavorable outcome. We also analyzed this dataset using logistic regression and found similar results (Supplemental Table 2).

**Table 2.** Primary (Arm B vs C) and secondary endpoints analyses. Univariate and multivariate analysis of variables associated with unfavorable outcomes combining Arms B and C (first column), Arms A + B + C (second column), or within Arm C alone (third column). Hazard ratios (HRs) > 1 indicate that increased values of the given characteristic correspond to faster times to relapse. Adjustments for multiplicity were not performed; p-values< 0.01 are in bold.

| Characteristic | B+C |  |  | A+B+C |  |  | C only |  |  |
| --- | --- | --- | --- | --- | --- | --- | --- | --- | --- |
|  | Univariate Cox PH Model |  |  | Univariate Cox PH Model |  |  | Univariate Cox PH Model |  |  |
|  | HR<sup>1</sup> | 95% CI<sup>1</sup> | p-value | HR<sup>1</sup> | 95% CI<sup>1</sup> | p-value | HR<sup>1</sup> | 95% CI<sup>1</sup> | p-value |
| <u>Arm</u> |  |  |  |  |  |  |  |  |  |
| B | — | — |  | — | — |  | — | — | — |
| C | 7.99 | 1.84, 34.7 | **0.006** | 8.19 | 1.89, 35.5 | **0.005** | — | — | — |
| A | — | — | — | 2.08 | 0.44, 9.81 | 0.4 | — | — | — |
| <u>Country</u> |  |  |  |  |  |  |  |  |  |
| CHN | — | — |  | — | — |  | — | — |  |
| RSA | 3.2 | 0.93, 11.0 | 0.064 | 4.08 | 1.23, 13.6 | 0.022 | 2.98 | 0.86, 10.4 | 0.086 |
| Age | 1 | 0.96, 1.03 | 0.9 | 1.01 | 0.98, 1.03 | 0.6 | 1.01 | 0.97, 1.04 | 0.7 |
| <u>Sex</u> |  |  |  |  |  |  |  |  |  |
| Female | — | — |  | — | — |  | — | — |  |
| Male | 1.26 | 0.45, 3.51 | 0.7 | 1.32 | 0.53, 3.28 | 0.5 | 1.53 | 0.50, 4.71 | 0.5 |
| <u>Prior TB episode</u> |  |  |  |  |  |  |  |  |  |
| Yes | — | — |  | — | — |  | — | — |  |
| No | 0.47 | 0.18, 1.25 | 0.13 | 0.49 | 0.22, 1.13 | 0.1 | 0.41 | 0.15, 1.11 | 0.08 |
| BMI | 0.84 | 0.70, 1.01 | 0.066 | 0.83 | 0.71, 0.98 | 0.023 | 0.83 | 0.68, 1.01 | 0.062 |
| Weight | 0.95 | 0.91, 1.00 | 0.068 | 0.95 | 0.91, 0.99 | 0.023 | 0.94 | 0.90, 1.00 | 0.039 |
| Number of cavities week 0 | 1.48 | 0.91, 2.41 | 0.12 | 1.33 | 0.96, 1.83 | 0.085 | 1.38 | 0.76, 2.51 | 0.3 |
| Cavity Volume week 0 (log2) | 1.07 | 0.98, 1.16 | 0.11 | 1.05 | 0.98, 1.12 | 0.2 | 1.06 | 0.97, 1.15 | 0.2 |
| Cavity Volume week 4 (log2) | 1.1 | 1.00, 1.21 | 0.063 | 1.05 | 0.97, 1.13 | 0.2 | 1.09 | 0.98, 1.21 | 0.11 |
| Cavity Volume week 16 (log2) | 1.19 | 1.08, 1.31 | **<0.001** | 1.11 | 1.02, 1.20 | 0.013 | 1.17 | 1.06, 1.30 | **0.002** |
| Cavity Volume change week 0 to 4 (log ratio) | 0.98 | 0.76, 1.26 | 0.9 | 0.94 | 0.75, 1.18 | 0.6 | 1 | 0.75, 1.35 | >0.9 |
| Cavity Volume change week 0 to 16 (log ratio) | 1.11 | 0.97, 1.27 | 0.13 | 1.08 | 0.96, 1.22 | 0.2 | 1.09 | 0.96, 1.24 | 0.2 |
| Cavity Volume change week 0 to 4 (percent) | 0.54 | 0.17, 1.66 | 0.3 | 0.44 | 0.17, 1.16 | 0.1 | 0.61 | 0.18, 2.04 | 0.4 |
| Cavity Volume change week 0 to 16 (percent) | 1.2 | 1.05, 1.38 | **0.008** | 1.19 | 1.02, 1.39 | 0.024 | 1.16 | 1.01, 1.33 | 0.039 |
| Total hard volume week 0 (log2) | 1.45 | 0.93, 2.26 | 0.1 | 1.17 | 0.87, 1.59 | 0.3 | 1.4 | 0.88, 2.22 | 0.2 |
| Total hard volume week 4 (log2) | 1.33 | 0.91, 1.95 | 0.14 | 1.06 | 0.82, 1.38 | 0.6 | 1.35 | 0.90, 2.04 | 0.15 |
| Total hard volume week 16 (log2) | 1.16 | 0.82, 1.65 | 0.4 | 1.09 | 0.82, 1.45 | 0.6 | 1.18 | 0.82, 1.72 | 0.4 |
| Hard Volume change week 0 to 4 (log ratio) | 0.96 | 0.45, 2.05 | >0.9 | 0.74 | 0.43, 1.27 | 0.3 | 1.14 | 0.53, 2.44 | 0.7 |
| Hard Volume change week 0 to 16 (log ratio) | 0.88 | 0.62, 1.24 | 0.5 | 0.93 | 0.66, 1.30 | 0.7 | 0.93 | 0.66, 1.32 | 0.7 |
| Hard Volume change week 0 to 4 (percent) | 0.91 | 0.16, 5.03 | >0.9 | 0.75 | 0.22, 2.59 | 0.6 | 1.25 | 0.24, 6.50 | 0.8 |
| Hard Volume change week 0 to 16 (percent) | 0.42 | 0.10, 1.69 | 0.2 | 0.54 | 0.16, 1.83 | 0.3 | 0.48 | 0.12, 1.89 | 0.3 |
| Total lesion glycolysis week 0 (log2) | 1.34 | 0.95, 1.89 | 0.1 | 1.17 | 0.89, 1.52 | 0.3 | 1.32 | 0.93, 1.88 | 0.12 |
| Total lesion glycolysis week 4 (log2) | 1.5 | 1.07, 2.11 | 0.018 | 1.21 | 0.95, 1.52 | 0.12 | 1.58 | 1.09, 2.28 | 0.015 |
| Total lesion glycolysis week 16 (log2) | 1.76 | 1.27, 2.45 | **<0.001** | 1.41 | 1.11, 1.80 | **0.005** | 1.97 | 1.32, 2.94 | **<0.001** |
| Total lesion glycolysis change week 0 to 4 (log ratio) | 1.47 | 0.96, 2.26 | 0.074 | 1.28 | 0.85, 1.93 | 0.2 | 1.5 | 0.99, 2.28 | 0.057 |
| Total lesion glycolysis change week 0 to 16 (log ratio) | 1.46 | 1.07, 1.98 | 0.015 | 1.46 | 1.09, 1.96 | 0.012 | 1.47 | 1.08, 2.00 | 0.014 |
| Total lesion glycolysis change week 0 to 4 (percent) | 1.04 | 0.69, 1.57 | 0.8 | 0.98 | 0.59, 1.64 | >0.9 | 1.04 | 0.71, 1.53 | 0.8 |
| Total lesion glycolysis change week 0 to 16 (percent) | 1.34 | 0.70, 2.58 | 0.4 | 1.43 | 0.79, 2.58 | 0.2 | 1.22 | 0.65, 2.32 | 0.5 |
| GeneXpert cycle threshold week 0 | 0.87 | 0.79, 0.97 | **0.009** | 0.92 | 0.84, 1.00 | 0.057 | 0.89 | 0.81, 0.98 | 0.016 |
| GeneXpert cycle threshold week 16 | 0.87 | 0.78, 0.97 | 0.012 | 1.01 | 0.95, 1.07 | 0.8 | 0.92 | 0.81, 1.05 | 0.2 |
| GeneXpert cycle threshold change week 0 to 16 | 1.05 | 0.97, 1.15 | 0.2 | 1.05 | 0.98, 1.11 | 0.15 | 1.08 | 0.99, 1.18 | 0.095 |
| MGIT time to detection week 0 (log2) | 0.89 | 0.48, 1.65 | 0.7 | 1.02 | 0.61, 1.70 | >0.9 | 1.07 | 0.55, 2.10 | 0.8 |
|  | **Multivariate Cox PH model** |  |  | **Multivariate Cox PH model** |  |  | **Multivariate Cox PH model** |  |  |
| <u>Arm</u> |  |  |  |  |  |  |  |  |  |
| B | — | — | — | — | — |  | — | — | — |
| C | 8.85 | 2.02, 38.8 | **0.004** | 10.6 | 2.39, 46.8 | **0.002** | — | — | — |
| A | — | — | — | 0.74 | 0.13, 4.25 | 0.7 | — | — | — |
| log2_CAVTOT_wk16 | 1.14 | 1.03, 1.26 | 0.014 | — | — | — | — | — | — |
| log2_TLG_wk16 | 1.71 | 1.17, 2.48 | **0.005** | 1.82 | 1.32, 2.49 | **<0.001** | 1.98 | 1.34, 2.92 | **<0.001** |
| BMI | — | — | — | 0.83 | 0.71, 0.98 | 0.024 | 0.83 | 0.69, 0.99 | 0.042 |

### Baseline cavities reactivate at relapse

PET/CT scans at time of TB recurrence were obtained in 21 of the 23 study participants who experienced confirmed relapse (Figure 2C). All participants who relapsed had cavities at baseline on PET/CT scan, although some were less than 2 ml in volume. In 17 of the 21 relapse scans there was disease reactivation by PET FDG-uptake in a cavity that was FDG-avid at the start of treatment and that had generally reduced uptake on treatment. Figure 3 shows two examples of relapse scans in Arm C participants. In the first (Figure 3A) a large left apical thick-walled cavity which appeared to be in close contact with the posterior pleural surface mostly resolved after four months of treatment. The residual lesions at four months appeared unremarkable but there was a strong pleural connection in the axial and sagittal sections. Treatment was complete at this time per protocol. Six months after completing treatment the same cavity recrudesced, and the pleural connection extended. In the second example (Figure 3B), the subject presented at baseline with bilateral apical disease with a large right apical cavity with evident liquified caseum (lack of FDG labeling in the interior region of the cavity, best seen as an air-fluid level in the axial section). This “dirty” cavity phenomenon was also evident in a left apical cavity which appeared to have only begun the process of liquefaction. The left apical cavity also appeared to show more intimate pleural adhesion than the right side. Both cavities appear to respond after four months of treatment although the left side lesion appeared to close off rather than resolve. Treatment completed at four months. Upon relapse, 14 months after therapy completion, only the left cavity appeared to be the major source of activity.

**Figure 3.**
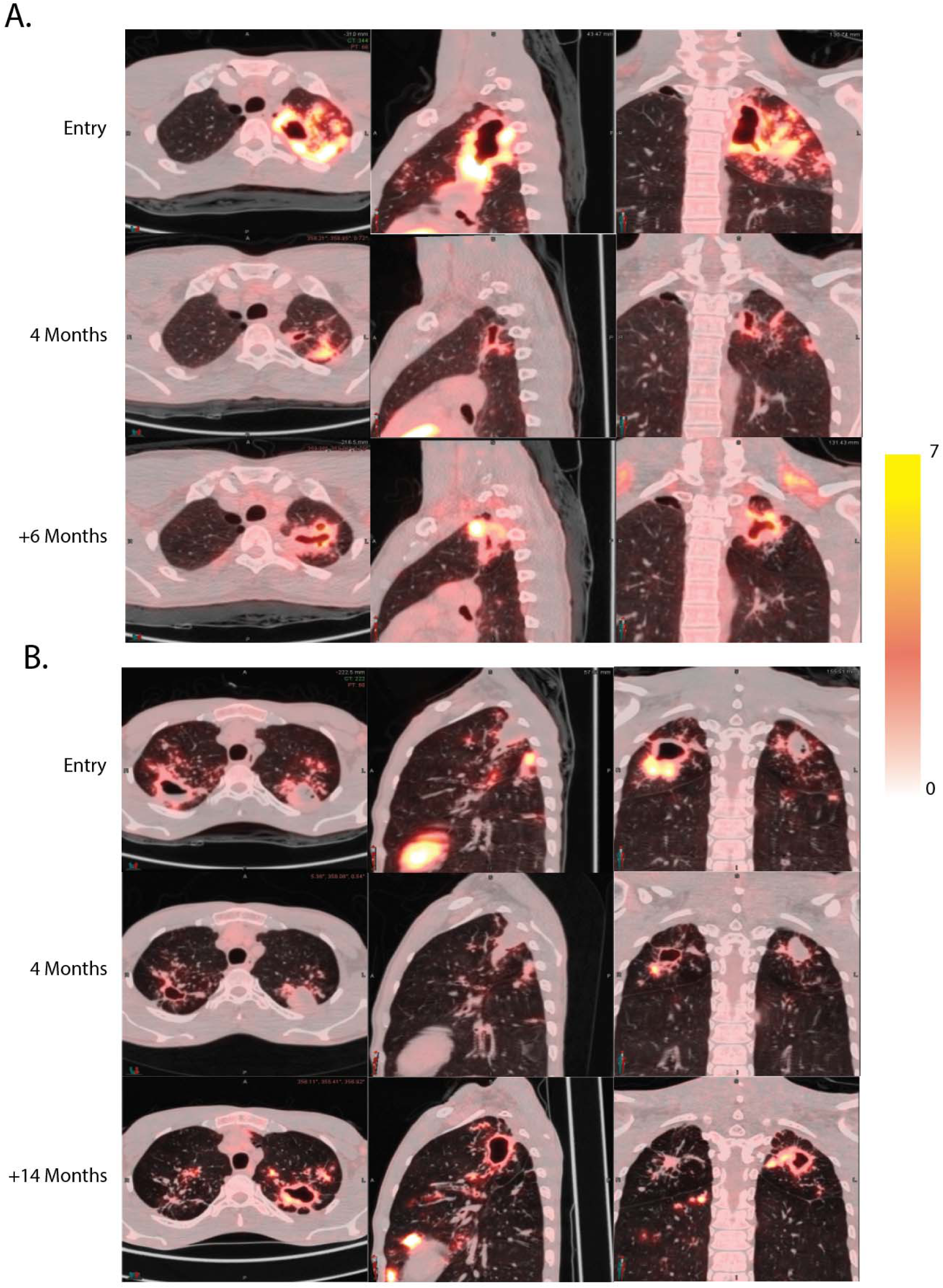
TB relapse involves baseline cavities. **A.** Subject A19 presented with a large left apical cavity which appeared mostly resolved after four months of treatment. Six months after completing therapy the participant became sputum culture positive again with the same isolate and with a large left apical cavity in the same region of the lung. **B.** Participant A9 presented with bilateral disease with a left upper “dirty” cavity with cold central necrotic material and only slight liquefaction. After four months of therapy the right apical cavity showed substantial response while the cold left side lesion appeared less active. 14 months later the participant experienced relapse and became sputum culture positive with an identical isolate and the necrotic left apical lesion appeared to be an active thick-walled cavity.

Two of the four participants who experienced confirmed relapse without reactivation of a cavitary lesion already present at baseline were related and cohabitated during the study and the timing of their diagnoses was possibly consistent with reinfection with the same strain (Supplemental Figure 2). In the third participant (A23), the relapse PET/CT scan showed a small cavity at the original cavity site, but a new posterior lesion connected to that cavity by an inflamed bronchus was the main active lesion (Supplemental Figure 3). As this scan was collected one year after treatment was discontinued, this new lesion was possibly seeded by the original cavity, a phenomenon we have previously reported(*20*). In the fourth participant (A27), there was no new lesion at the time of relapse and only sporadic low-level culture positivity, however this scan was obtained five weeks after the participant restarted therapy making it difficult to know what the appearance was when the participant was diagnosed with relapse.

In four participants with failed treatment (defined as relapse by week 24), the active lesions observed on their third (and final) scans were all in the same location as their baseline lesions. On recurrence scans of all three cases of reinfection, active lesions were observed in areas of the lung that did not show involvement on prior scans and lesions present at baseline did not appear active at recurrence (see Supplemental Figure 4 for an example).

### Computational extraction and analysis of primary statistics

To gain deeper insights into the features linked to relapse, we segmented the entire set of PET/CT scans using an enhanced version of our previously described automated computational method(*21, 22*). All scans were subjected to segmentation and then inspected manually (Figure 4). High quality masks were produced from the CT scans and then subjected to rigid co-registration based on the carina across visits. Sub-segmentations (masks) were also created for cavities (any interior airspace with contiguous lesion voxels around the air to capture cavity walls), hard lesion volume (HU values between -100 and +200) and soft lesion volume (abnormalities with HU density -500 to -101). Each voxel within a lesion mask was then used to compute various statistics of the material in the CT scan such as average, max and min HU (a full list of computed variables and an example are shown in Supplemental Methods S1.1). Co-registered PET scans have larger voxel sizes, so PET statistics were calculated and then imputed to all CT voxels contained within each PET voxel. This refinement includes the introduction of the Relapse Active Zone (RAZ) as detailed in Supplemental Methods S1.1. Briefly all scans were scaled to the same size and a common coordinate system within which comparable volumes could be compared and those regions and their features associated with relapse were identified.

**Figure 4.**
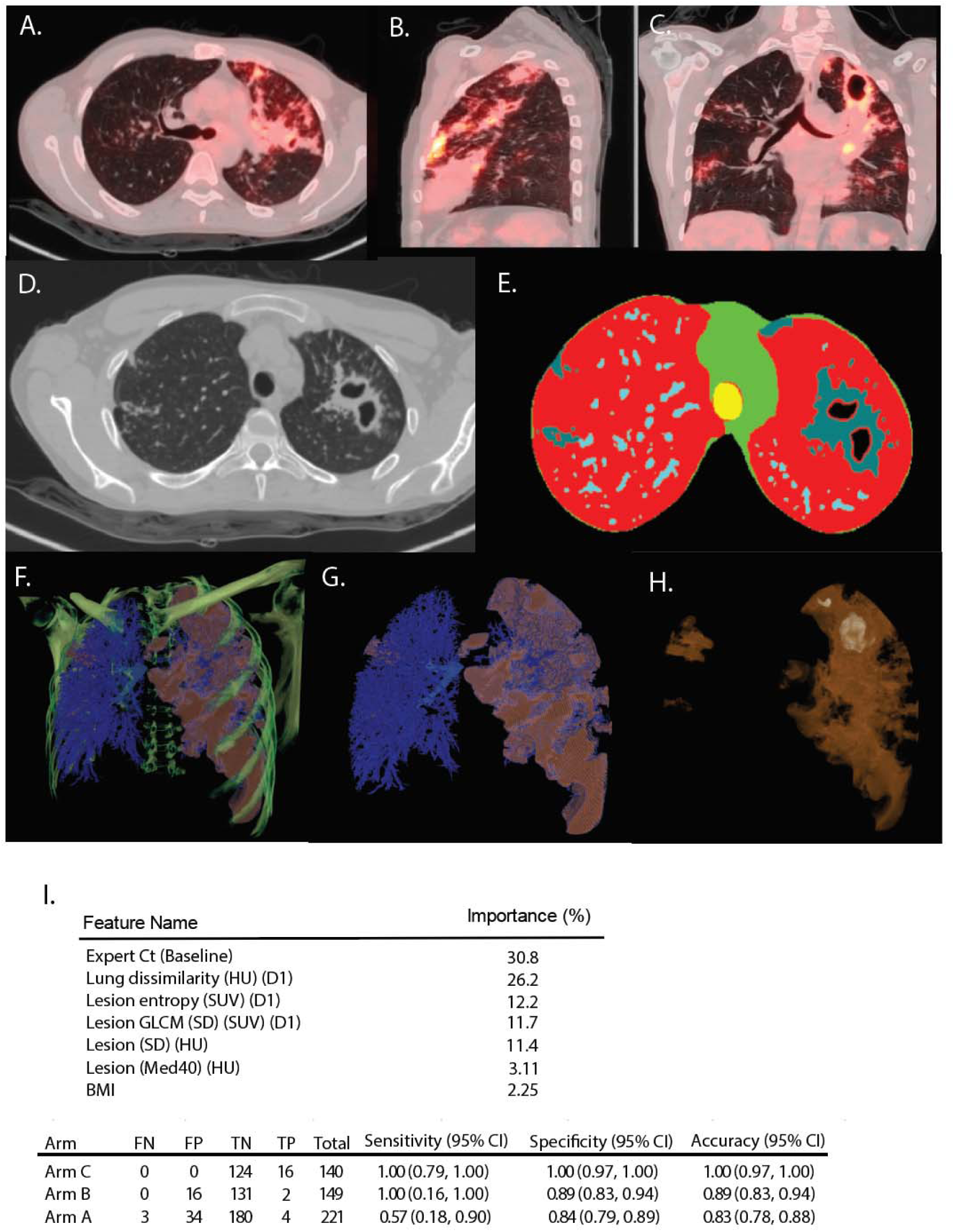
Segmentation and computational scan analysis. (**A**) Axial, (**B**) Sagittal, (**C**) Coronal views of PET/CT scan of subject A1. CT scans viewed in lung window and PET at an SUV of 7 are displayed. (**D**) CT slice from A1 showing cavity and associated features in a single slice, (E) segmented mask for the same CT slice as shown in **D**, low density normal lung voxels colored red, high density mediastinal and surface of thoracic cavity shown in green, trachea colored yellow, pulmonary vasculature shown in light blue and TB-associated lesional material shown in grey, (**F**) 3D multiplanar view of the same scan showing the CT bone density colored in green superimposed on a rendering of the 3D masks (low density normal lung removed for clarity) vasculature mask and major airways shown in blue, lesion material shown in peach. (**G**) 3D rendering of the vascular and lesion masks of A1 colored as in **F** with the primary CT data including the bones removed. (**H**) 3D rendering of only the lesion material in A1 with enhanced transparency to view the large left apical cavity. (**I**) Machine learning model for relapse prediction using Arm C PET/CT scans as the training set and Arms A and B PET/CT scans as the test set. Significant features are Xpert Ct (Baseline): GeneXpert cycle threshold value at baseline, Lung Dissimilarity (HU) (D1): Dissimilarity in Hounsfield Units (HU) between adjacent voxels at a distance of 1 voxel, Lesion entropy (SUV) (D1): Entropy calculated across a lesion at a distance of 1 voxel, Lesion GLCM (gray level co-occurrence matrix) (SD)(SUV)(D1): Standard deviation of the Standardized Uptake Value (SUV) calculated across the lesion at a distance of 1 voxel, Lesion (SD) (HU): standard deviation of HU within a lesion, Lesion (MED40) (HU): Lesion level 40^th^ percentile of HU value, Lesion Energy (HU) (D1): Energy computed on HU values across a lesion at a distance of 1 voxel, BMI: Body Mass Index.

Segmentation was further refined by associating physically connected voxels in two categories: first for “lesions” without interior air spaces and second for “cavities” – lesions with interior air spaces. Participants contained an average of 51 lesions (range 1 – 664, Std Dev 122) and 6 cavities (range 1-134, Std Dev 16), although many of these features were small (of 3,385 cavities in the total baseline dataset, 2650 (78.3%) were smaller than 1 mL). The same segmentation process was applied to all scans during the study and co-registered coordinates used to compute changes in individual lesions over time. For each lesion we calculated first-order attributes for these subsegments including voxel count, volume, minimum and maximum HU and SUV values, standard deviations of voxel values and a variety of other values (Full list in Supplemental Methods, Table S1.4). We also calculated a variety of texture attributes including contrast, dissimilarity, homogeneity, energy, entropy and grey-level correlation metrics (full list in Supplemental Methods, Table S1.1).

Using the full set of both PET and CT statistics from participants in Arm C (at baseline), we trained a machine learning algorithm (Supplemental Methods S1.2) to discriminate relapse from cured participants. Based on the top three performing configurations, the algorithm demonstrated excellent performance on the training set (Arm C), achieving zero false negatives and false positives in this cohort. However, this perfect performance suggests overfitting, meaning the model may have learned to recognize patterns specific to the training data that do not generalize well to new, more diverse patient populations with varying disease characteristics and treatment regimens. As expected, the model’s performance was reduced when applied to the test cohorts, where patient characteristics and treatment regimens differed from those in the training set, but still performed reasonably well. When applied separately to the test cohorts from Arms A and B, where the treatment duration was six months, the model’s accuracy remained high, with scores of 0.83 (95% CI: 0.78, 0.88) and 0.89 (95% CI: 0.83, 0.94), respectively.

In Arm A, the model’s sensitivity dropped to 0.57 (95% CI: 0.18, 0.90). This decline can be attributed to two main factors. First, Arm A had a higher proportion of patients with large cavities and large consolidations, compared to Arm C, on which the model was trained. Participants with advanced disease, characterized by larger cavitary lesions, more extensive lung involvement, and greater disease burden, were by design stratified to Arm A and the model was not developed on these advanced disease features. These results highlight the model’s strengths in identifying ‘easy to treat’ patients, particularly those with less severe disease profiles and more predictable responses to treatment, providing a valuable foundation for further optimization.

In contrast, the model performed exceptionally well in Arm B, maintaining a sensitivity of 1.00 (95% CI: 0.16, 1.0). This high sensitivity is likely because Arm B participants had similar baseline disease burden and early treatment response as Arm C participants. The wide confidence interval reflects the uncertainty in this estimate due to the few (2) poor outcomes. The model’s robust performance in Arm B suggests its potential utility in identifying relapse risk in patients with less severe disease profiles, where clinical characteristics closely align with the training data, even when the treatment duration is extended.

## Discussion

Our study hypothesized that: 1) TB disease severity at baseline by PET/CT criteria; 2) early treatment response, based on PET/CT criteria at one month, with an emphasis on cavities, TLG and hard disease volume; and 3) a GeneXpert cycle threshold minimum at four months, could stratify patients into two cohorts, one less severely diseased and eligible for treatment shortening, and the other more severely diseased and not eligible for treatment shortening. We based these criteria on a smaller cohort of participants with PET/CT data available(*23*). These criteria were clearly unsuccessful at stratifying easier-to-treat versus harder-to-treat participants. Our unfavorable outcome rates in the two six-month arms were 3.2% (Arm A) and 1.5% (Arm B) while the rate of unfavorable outcomes was 12.1% in the four-month treatment arm (Arm C). These outcomes were consistent with those observed in other studies which did not stratify participants by disease burden and response, ReMoxTB, for example, which had 2% relapse in the six-month control arm (and 3% retreatment) and 9-12% relapse in both four-month treatment arms(*16, 19*).

Our first stratification criterion was no single large cavitary lesion (>30mL) visible in the baseline CT scan. Cavities of this size are relatively uncommon and only 30 participants in the entire cohort had such large cavities (Supplemental Figure 5A). All of these were assigned by protocol to Arm A and three of these participants experienced an unfavorable outcome (2 relapse, 1 treatment failure). Amongst the unfavorable outcomes in Arm C that were not reinfection, the cavities that appeared to cause relapse ranged from 1-15mL in interior air volume (Supplemental Figure 5B). Often these cavities appeared to be early in the process of liquefaction with evident caseum and thick walls and they frequently had intimate contact with the pleural space. Analyses of these particular features and how they correlate with treatment outcomes are ongoing.

Our second stratification criterion was related to the total hard CT volume or total PET activity at baseline. Amongst the participants assigned to Arm A and designated harder to treat, these two features accounted for the reason in 48% of the total while large cavities only accounted for 13% (Supplemental Figure 1). Hard CT volume represents a complex mixture of pathologies in TB disease ranging from cavity walls to larger areas of inflammation, fibrosis and dense airway infiltrates. Neither criterion was found to be significantly associated with outcome in this study. Instead, the sixteen-week total glycolytic activity was significant in both univariate and multivariate analyses. This suggests that some lesions with significant contributions to hard volume and total glycolytic activity are present at baseline but do not contribute to outcome prognosis – either because they are easier to treat or fibrotic. During the conduct of this study, we noted new lesions associated with cavities through bronchial spread(*20*). Such newly formed infiltrates are an obvious candidate for lesions that resolve quickly as they lack fibrotic structures that surround cavities and large nodules that limit drug access(*24*). These infiltrates are an important contributor to total glycolytic activity at baseline and early treatment and appear to respond relatively quickly to treatment.

Our third criterion referenced the improvement observed between the baseline and the week 4 PET/CT scans. In this study reduction in cavity volume at week 16 was significant in univariate but not in the multivariate analysis. Likewise, CT hard volume and PET activity reductions at week 4 were not significantly associated with relapse. In the end, arm A (participants meeting our definition of having higher risk for relapse) had only a very slight enrichment for relapse compared to Arm B (3.2% vs 1.5%) suggesting that our criteria based on the previous study did not identify patients at higher risk for relapse. The one-month timing of this evaluation was probably confounded by flares in glycolytic activity in many participants who otherwise did well. These flares occurred in pre-existing lesions and were distinct from the appearance of new airway lesions described above. From the computationally segmented lungs, we observed that nearly one-third of all scanned participants (202 out of 648, 31%) experienced an increase in TGA in their largest lesion at one month (Supplemental Figure 6). This is likely a manifestation of a subclinical paradoxical reaction which is common even in HIV-uninfected individuals(*25*). Paradoxical reactions are more frequently reported in extrapulmonary tuberculosis(*26*) although it may be that sequalae of such cases are more pronounced and it is therefore detected more frequently than in patients with primarily pulmonary disease.

Our study did establish unambiguously that cavities already present at baseline played a central role in relapse. This is consistent with the significant association seen with baseline cavity volume in our, and many other(*5-9*), studies. In addition, we noted that cavitary lesions with thick walls were present in nearly all relapse cases although they were also present in a large fraction of participants who did not relapse. The liquifying caseum within a thick-walled cavity was first noted as early as 1946 by the pioneering work of Canetti(*27*) who noted that the interior surface of the cavity was a primary site of rapid bacillary expansion and that as one proceeded towards the capsular lining the material was increasingly paucibacillary caseum. These “dirty” cavities are transient and ultimately result in a fibrous shell, Canetti proposed that such active processes are often over in a matter of days, a proposal supported by the rapidity of liquefaction in non-human primates(*28*). It seems possible that the application of chemotherapy during liquefaction interrupts the process to leave a caseum-adapted population of bacilli poised to reactivate when the pressure is removed.

To understand why some cavities are more difficult to treat than others we computationally segmented all PET/CT scans obtained during this study. This allowed us to compute detailed statistics for large numbers of lesions from each subject at multiple time points and apply machine learning to predict features that were associated with relapse. The resulting model was highly successful at identifying lesions associated with relapse, perfectly in Arm C patients used in training the model but also performed well in Arm B participants, which the model had never seen. Performance of the model in the non-randomized Arm A patients was lower. Our data represent the largest known set of TB-relapses characterized by PET-CT imaging. The model could be improved by including more participants with extensive disease like that seen in Arm A. Evaluation of model performance in an independent population of TB patients is warranted. Three textural features account for almost 50% of the model’s predictive ability including lesion dissimilarity in radiodensity, lesion entropy in SUV and the correlation of SUV values in a grey layer correlation matrix. The pathophysiology resulting in these textural features are not intuitively obvious but speak to a heterogenous distribution of values for SUV and HU across lesions that place a subject at higher risk for relapse with early treatment discontinuation. One unappreciated feature that could contribute to such heterogeneity is the extent of pleural adhesions of cavities to the chest wall. From the pre-chemotherapy era we know that pleural adhesions are very common in autopsies of people who died from tuberculosis. For example, in a review of 505 autopsies of tuberculosis patients in Los Angeles from 1911-1950 as many as 85% displayed pleural adhesions(*29*). From the more modern era pleural adhesions have been described primarily as complications of surgical interventions to remove infected lungs(*30*). Interestingly the main surgical complications appear to result from excess bleeding since these adhesions can recruit blood vessels from outside the thoracic cavity directly to the lesion.

Recent reanalysis of data from trials including the failed 4-month treatment shortening trials(*16-18*) has used some simple clinical features to stratify TB patients at baseline into three categories; low, moderate and high risk for relapse(*31*). Using this model the low-risk group was shown to be non-inferior to standard of care after four months. The failure to achieve non-inferiority in the full cohort was entirely due to the participants in the moderate and high-risk categories. Although this model was good, our machine learning model based on both clinical and radiological features may be better to stratify participants in future trials to increase the representation of participants with harder-to-treat lesions. This could reduce the size of future studies significantly and allow more cost-effective comparisons of different regimens. For example, the model based on clinical features could be used to screen for participants that would receive the more detailed radiology studies necessary to exclusively enroll “harder to treat” patients with a high likelihood of relapse. This would allow a comparison of the treatment-shortening potential of multiple regimens in Phase 2 prior to embarking on large Phase 3 studies. Alternatively, the model could enable testing of only low risk patients to develop regimens that could result in much shorter regimens for TB patients with low-risk disease.

Aside from the utility of the models in trials, our results suggest an explanation for the unusual distribution of relapses observed across all TB patients where 80% of patients are durably cured after 3-4 months of therapy: relapses result from cavitary lesions with specific radiologic features that are not present in most newly diagnosed patients. This implies that the bacilli within these lesions are either fundamentally different from (e.g. sub-breakpoint MIC) or harder to treat than (e.g. due to drug penetration) other pathological manifestations and this finding will be helpful in focusing the search for new antitubercular medications active against these bacilli, allowing treatment shortening for even patients currently considered “high-risk”.

## MATERIALS AND METHODS

“Using biomarkers to predict TB treatment duration (PredictTB)” was a prospective, multicenter, non-inferiority, treatment-shortening, randomized clinical trial of adult pulmonary TB patients that has been previously described(*23*). Briefly, adult pulmonary TB patients were enrolled from 5 sites in the Western Cape, South Africa (Stellenbosch University, University of Cape Town Lung Institute, University of Cape Town South African Tuberculosis Vaccine Initiative, Khayelitsha Site B, and TASK) and 5 sites in Henan Province, China (Henan Provincial Chest Hospital, Xinmi City Institute of Tuberculosis Prevention and Control, Kaifeng City Institute of Tuberculosis Prevention and Control, Zhongmu County Health and Epidemic Prevention Station, and Xinxiang City Institute of Tuberculosis Prevention and Control). Inclusion criteria included adult pulmonary TB patients who were GeneXpert MTB/RIF positive for TB and rifampin sensitive who had not yet started TB treatment. Exclusion criteria included extrapulmonary TB, pregnancy, HIV positivity, diabetes, INH mono-resistance, or use of any immunosuppressive medications. Eligible participants signed informed consent and were started on standard of care fixed dose combination TB treatment with isoniazid, rifampicin, pyrazinamide, and ethambutol for 8 weeks, followed by isoniazid and rifampicin until treatment completion.

All participants received a thoracic FDG PET/CT scan at baseline and week 4. Participants in Arm B and C were all scanned at week 16 as well, and participants in Arm A randomly had their third scan at either week 16 or 24. A possible 4^th^ scan was done at the time of TB recurrence. A pre-specified scan reading algorithm was developed based on data from an earlier study(*32*) and two trained scan readers were randomly selected from a team of twelve physicians to read each scan. Scans with discrepant arm assignments between readers were read by a third reader. Participants were considered “low risk” if their baseline PET/CT scans did not show the following: total collapse of a single lung, a single cavity with a volume >30mL, CT scan hard volume (-100 to +100HU density) <200 mL or PET total glycolytic activity <1500 units. Participants who exceeded any of these criteria were assigned to Arm A and received six months of standard treatment while participants who met all of these criteria were assigned to Arms B/C and started on standard treatment until one month when they received a repeat PET/CT scan. At the week 4 PET/CT scan, if participants failed to show a 20% decrease in cavitary air volume, showed an increase in hard volume of more than 10%, or an increase in PET glycolytic activity of more than 30% they were assigned to Arm A and not eligible for randomization. At week 16, participants still in Arms B/C were moved to Arm A if a GeneXpert cycle threshold was <30 (criterion changed from <28 after the initial 10 participants were randomized, due to more participants than expected *a priori* being stratified to Arm A at week 16) or if the participant had not taken at least 100/112 doses, based on data from the Medication Event Reminder Monitor (MERM), an electronic pill box(*33, 34*). Participants stratified to Arm A at baseline, week 4, or week 16 received standard of care TB treatment for 24 weeks. Participants who met all treatment shortening criteria at all 3 timepoints were randomized to Arm B and received standard of care TB treatment for 24 weeks, or to Arm C and completed treatment at 16 weeks. All participants were followed to 72 weeks for final treatment outcomes. Patients who developed recurrent TB during follow-up had repeat PET/CT exams and their bacterial strains were isolated and whole genome sequencing was performed to distinguish relapse from reinfection.

Informed consent was obtained in the local language of the participant (English, Afrikaans, Xhosa, or Chinese). This study was reviewed and approved by the IRB/ethics committees of the NIAID, Stellenbosch University, Faculty of Health Sciences University of Cape Town, South African Medicine Control Council, Henan Provincial Chest Hospital, and the Henan Center for Disease Control. The standing NIAID DSMB with three global TB experts added as *ad hoc* members provided oversight of the trial, meeting at least twice per year to review study data. Pre-specified stopping rules for inferiority of the treatment shortening arm and for study futility were provided to the DSMB. This study is registered at www.clinicaltrials.gov (NCT02821832). The first participant was enrolled in June 2017 in the Western Cape and in October 2017 in Henan Province. The last participant was enrolled in September 2020 in Henan and in March 2021 in the Western Cape, with follow-up completing 18 months later.

### Statistical methods

Cox proportional hazards models for the time to relapse/poor outcomes were fit in univariate and multivariate analyses to evaluate the associations between PET/CT characteristics, GeneXpert results, and host factors. Logistic regression analyses were fit for comparisons to Cox proportional hazards model. Confidence intervals for diagnostic accuracy, sensitivity and specificity were estimated using binomial exact confidence intervals. Analyses were conducted in R.

## Data availability statement

All data related to the conduct of this trial are maintained in a locked, secure database and will be made available to any qualified investigator upon completion of a clinical data sharing agreement with NIAID.

## Acknowledgements

We thank all individuals that participated in this study, and all staff at the various recruitment sites mentioned above. We also acknowledge the Western Cape Academic PET/CT Centre, Numeri Node for Infectious Imaging, Cape Universities Body Imaging Centre (CUBIC), Cape PET/CT Centre, Henan People’s Hospital, and the Henan Cancer Hospital for access to PET-CT scans, as well as Ithemba Laboratories and PET LABS for FDG production, and Cepheid for providing GeneXpert instruments. We thank all clinical and laboratory staff employed by the Centre for Infectious Diseases Research in Africa for their contribution to the study. Further to the Western Cape Department of Health and Wellness, and City of Cape Town Health for facilitating access to facilities to conduct the study.

