## Supplemental Information for "PET/CT guided tuberculosis treatment shortening: a randomized trial"

Supplemental Table 1. Reasons for exclusion from study

| Criteria | # Excluded |
|---|---|
| 1) Clinical suspicion of or confirmed extrapulmonary TB, including pleural TB | 10 |
| 2) Pregnant or desiring/trying to become pregnant in the next 6 months or breastfeeding | 6 |
| 3) HIV infected | 17 |
| 4) Currently COVID-19 infected | 2 |
| 5) Unable to take oral medications | 0 |
| 6) Diabetes as defined by point of care HbA1c ≥6.5%, random glucose ≥200 mg/dL (or 11.1 mmol/L), fasting plasma glucose ≥126 mg/dL (or 7.0 mmol/L), or the presence of any anti-diabetic agent (including traditional medicines) as a concomitant medicine | 43 |
| 7) Disease complications or concomitant illnesses that may compromise safety or interpretation of trial endpoints, such as known diagnosis of chronic inflammatory condition (e.g. sarcoidosis, rheumatoid arthritis, connective tissue disorder) | 1 |
| 8) Use of immunosuppressive medications, such as TNF-alpha inhibitors or systemic or inhaled corticosteroids, within the past 2 weeks | 13 |
| 9) Use of any investigational drug in the previous 3 months | 1 |
| 10) Substance or alcohol abuse that in the opinion of the investigator may interfere with the participant's adherence to study procedures | 2 |
| 11) Any person for whom the physician feels this study is not appropriate | 2 |
| 12) Not met inclusion criteria (Xpert-, low weight, RIF-R, no sputum production, Blood chemistry (AST ALT HB), BL scan unavailable) | 171 |
| Sum: | 268 |

Supplemental Table 2A : Additional statistical analyses of all three arms.

The relationship between demographic, clinical, and laboratory biomarkers and treatment outcomes. We used univariate Cox proportional hazard models to identify significant biomarkers (p-value < 0.05), which were then included in a step-wise backward selection process. In this process, the covariate with the largest p-value was removed one at a time until all remaining p-values were less than 0.05. Additionally, we constructed logistic regression models using the same procedure to validate the results from the Cox PH.

### A + B + C, univariate

| | Cox PH Model | | | Logistic Model | | |
|---|---|---|---|---|---|---|
| **Characteristic** | **HR**[1] | **95% CI**[1] | **p-value** | **OR**[1] | **95% CI**[1] | **p-value** |
| **arm** | | | | | | |
| B | — | — | | — | — | |
| C | 8.19 | 1.89, 35.5 | 0.005 | 8.79 | 2.45, 56.2 | 0.004 |
| A | 2.08 | 0.44, 9.81 | 0.4 | 2.15 | 0.53, 14.4 | 0.3 |
| **country** | | | | | | |
| CHN | — | — | | — | — | |
| RSA | 4.08 | 1.23, 13.6 | 0.022 | 4.35 | 1.49, 18.5 | 0.018 |
| **Cavity Air (log2, wk0)** | 1.05 | 0.98, 1.12 | 0.2 | 1.05 | 0.98, 1.13 | 0.2 |
| **Cavity Air (log2, wk4)** | 1.05 | 0.97, 1.13 | 0.2 | 1.05 | 0.98, 1.14 | 0.2 |
| **Cavity Air (log2, wk16)** | 1.11 | 1.02, 1.20 | 0.013 | 1.12 | 1.03, 1.22 | 0.009 |
| **Cavity Air Ratio (log 2, wk0/wk4)** | 0.94 | 0.75, 1.18 | 0.6 | 0.93 | 0.75, 1.20 | 0.5 |
| **Cavity Air Ratio (log2, wk0/wk16)** | 1.08 | 0.96, 1.22 | 0.2 | 1.09 | 0.97, 1.24 | 0.2 |
| **Cavity Air Percent Change (wk0-wk4)** | 0.44 | 0.17, 1.16 | 0.10 | 0.42 | 0.15, 1.14 | 0.090 |
| **Cavity Air Percent Change (wk0-wk16)** | 1.19 | 1.02, 1.39 | 0.024 | 1.23 | 0.96, 1.65 | 0.080 |
| **CT Hard Volume (log2, wk0)** | 1.17 | 0.87, 1.59 | 0.3 | 1.19 | 0.88, 1.66 | 0.3 |
| **CT Hard Volume (log2, wk4)** | 1.06 | 0.82, 1.38 | 0.6 | 1.08 | 0.84, 1.42 | 0.6 |
| **CT Hard Volume (log2, wk16)** | 1.09 | 0.82, 1.45 | 0.6 | 1.11 | 0.84, 1.51 | 0.5 |
| **CT Hard Volume Ratio (wk0/wk4, log2)** | 0.74 | 0.43, 1.27 | 0.3 | 0.76 | 0.45, 1.37 | 0.3 |
| **CT Hard Volume Ratio (wk0/wk16, log 2)** | 0.93 | 0.66, 1.30 | 0.7 | 0.92 | 0.66, 1.33 | 0.7 |
| **CT Hard Volume Percent Change (wk0-wk4)** | 0.75 | 0.22, 2.59 | 0.6 | 0.78 | 0.21, 2.65 | 0.7 |
| **CT Hard Volume Percent Change (wk0-wk16)** | 0.54 | 0.16, 1.83 | 0.3 | 0.53 | 0.13, 1.52 | 0.3 |
| **Total Lesion Glycolysis (wk0, log2)** | 1.17 | 0.89, 1.52 | 0.3 | 1.18 | 0.91, 1.58 | 0.2 |
| **Total Lesion Glycolysis (wk4, log2)** | 1.21 | 0.95, 1.52 | 0.12 | 1.22 | 0.97, 1.58 | 0.11 |
| **Total Lesion Glycolysis (wk16, log2)** | 1.41 | 1.11, 1.80 | 0.005 | 1.45 | 1.14, 1.90 | 0.004 |
| **Total Lesion Glycolysis Ratio (wk0/wk4, log 2)** | 1.28 | 0.85, 1.93 | 0.2 | 1.31 | 0.86, 2.03 | 0.2 |

| | | | | | | |
|---|---|---|---|---|---|---|
| **Total Lesion Glycolysis Ratio (wk0/wk16, log 2)** | 1.46 | 1.09, 1.96 | 0.012 | 1.49 | 1.11, 2.08 | 0.013 |
| **Total Lesion Glycolysis Percent Change (wk0/wk4)** | 0.98 | 0.59, 1.64 | >0.9 | 1.0 | 0.44, 1.41 | >0.9 |
| **Total Lesion Glycolysis Percent Change (wk0/w16)** | 1.43 | 0.79, 2.58 | 0.2 | 1.49 | 0.55, 3.17 | 0.3 |
| **GeneXpert Cycle Threshold (wk0)** | 0.92 | 0.84, 1.00 | 0.057 | 0.92 | 0.83, 1.00 | 0.054 |
| **GeneXpert Cycle Threshold (wk16)** | 1.01 | 0.95, 1.07 | 0.8 | 1.01 | 0.95, 1.08 | 0.7 |
| **GeneXpert Cycle Threshold Change (wk0-wk16)** | 1.05 | 0.98, 1.11 | 0.15 | 1.05 | 0.99, 1.12 | 0.12 |
| **MGIT TTD (log2)** | 1.02 | 0.61, 1.70 | >0.9 | 1.02 | 0.60, 1.70 | >0.9 |
| **Age** | 1.01 | 0.98, 1.03 | 0.6 | 1.01 | 0.98, 1.03 | 0.7 |
| **Sex** | | | | | | |
| Female | — | — | | — | — | |
| Male | 1.32 | 0.53, 3.28 | 0.5 | 1.38 | 0.58, 3.83 | 0.5 |
| **Prior TB** | | | | | | |
| Yes | — | — | | — | — | |
| No | 0.49 | 0.22, 1.13 | 0.10 | 0.51 | 0.22, 1.27 | 0.12 |
| **BMI** | 0.83 | 0.71, 0.98 | 0.023 | 0.83 | 0.70, 0.96 | 0.022 |
| **Weight** | 0.95 | 0.91, 0.99 | 0.023 | 0.95 | 0.91, 0.99 | 0.024 |
| **Cavity Present (wk0)** | | | | | | |
| No | — | — | | — | — | |
| Yes | 1.75 | 0.77, 4.01 | 0.2 | 1.82 | 0.81, 4.50 | 0.2 |
| **Cavity Number wk0** | 1.33 | 0.96, 1.83 | 0.085 | 1.33 | 0.93, 1.84 | 0.093 |

[1]HR = Hazard Ratio. CI = Confidence Interval. OR = Odds Ratio

**A+B+C, multivariate**

**Cox PH regression results:**

| **Characteristic** | **HR**[1] | **95% CI**[1] | **p-value** |
|---|---|---|---|
| Arm | | | |
| B | — | — | |
| C | 10.6 | 2.39, 46.8 | 0.002 |
| A | 0.74 | 0.13, 4.25 | 0.7 |
| Tot. Les. Gly. (log2, wk16) | 1.82 | 1.32, 2.49 | <0.001 |
| BMI | 0.83 | 0.71, 0.98 | 0.024 |

[1]HR = Hazard Ratio. CI = Confidence Interval

**Logistic regression results:**

| **Characteristic** | **OR**[1] | **95% CI**[1] | **p-value** |
|---|---|---|---|
| Arm | | | |
| B | — | — | |
| C | 13.3 | 3.36, 91.1 | 0.001 |
| A | 0.69 | 0.12, 5.48 | 0.7 |
| Tot. Les. Gly. (log2, wk16) | 1.90 | 1.38, 2.77 | <0.001 |
| BMI | 0.79 | 0.64, 0.95 | 0.019 |

[1]OR = Odds Ratio. CI = Confidence Interval

Supplemental Table 2B : Additional statistical analyses of both randomized arms.

### B + C, univariate

| | Cox PH Model | | | Logistic Model | | |
|---|---|---|---|---|---|---|
| **Characteristic** | **HR[1]** | **95% CI[1]** | **p-value** | **OR[1]** | **95% CI[1]** | **p-value** |
| **arm** | | | | | | |
| B | — | — | | — | — | |
| C | 7.99 | 1.84, 34.7 | 0.006 | 8.79 | 2.45, 56.2 | 0.004 |
| **country** | | | | | | |
| CHN | — | — | | — | — | |
| RSA | 3.20 | 0.93, 11.0 | 0.064 | 3.42 | 1.10, 15.0 | 0.056 |
| **Cavity Air (log2, wk0)** | 1.07 | 0.98, 1.16 | 0.11 | 1.07 | 0.99, 1.17 | 0.10 |
| **Cavity Air (log2, wk4)** | 1.10 | 1.00, 1.21 | 0.063 | 1.10 | 1.00, 1.22 | 0.054 |
| **Cavity Air (log2, wk16)** | 1.19 | 1.08, 1.31 | <0.001 | 1.21 | 1.09, 1.35 | <0.001 |
| **Cavity Air Ratio (log 2, wk0/wk4)** | 0.98 | 0.76, 1.26 | 0.9 | 0.98 | 0.77, 1.30 | 0.9 |
| **Cavity Air Ratio (log2, wk0/wk16)** | 1.11 | 0.97, 1.27 | 0.13 | 1.12 | 0.98, 1.30 | 0.12 |
| **Cavity Air Percent Change (wk0-wk4)** | 0.54 | 0.17, 1.66 | 0.3 | 0.50 | 0.15, 1.67 | 0.3 |
| **Cavity Air Percent Change (wk0-wk16)** | 1.20 | 1.05, 1.38 | 0.008 | 1.30 | 1.00, 2.31 | 0.10 |
| **CT Hard Volume (log2, wk0)** | 1.45 | 0.93, 2.26 | 0.10 | 1.52 | 0.99, 2.52 | 0.081 |
| **CT Hard Volume (log2, wk4)** | 1.33 | 0.91, 1.95 | 0.14 | 1.40 | 0.96, 2.16 | 0.10 |
| **CT Hard Volume (log2, wk16)** | 1.16 | 0.82, 1.65 | 0.4 | 1.20 | 0.85, 1.76 | 0.3 |
| **CT Hard Volume Ratio (wk0/wk4, log2)** | 0.96 | 0.45, 2.05 | >0.9 | 1.03 | 0.49, 2.38 | >0.9 |
| **CT Hard Volume Ratio (wk0/wk16, log 2)** | 0.88 | 0.62, 1.24 | 0.5 | 0.87 | 0.61, 1.27 | 0.5 |
| **CT Hard Volume Percent Change (wk0-wk4)** | 0.91 | 0.16, 5.03 | >0.9 | 1.04 | 0.18, 5.69 | >0.9 |
| **CT Hard Volume Percent Change (wk0-wk16)** | 0.42 | 0.10, 1.69 | 0.2 | 0.42 | 0.08, 1.36 | 0.2 |
| **Total Lesion Glycolysis (wk0, log2)** | 1.34 | 0.95, 1.89 | 0.10 | 1.37 | 0.99, 2.03 | 0.082 |
| **Total Lesion Glycolysis (wk4, log2)** | 1.50 | 1.07, 2.11 | 0.018 | 1.54 | 1.11, 2.26 | 0.018 |
| **Total Lesion Glycolysis (wk16, log2)** | 1.76 | 1.27, 2.45 | <0.001 | 1.87 | 1.34, 2.78 | <0.001 |
| **Total Lesion Glycolysis Ratio (wk0/wk4, log 2)** | 1.47 | 0.96, 2.26 | 0.074 | 1.52 | 0.94, 2.48 | 0.088 |
| **Total Lesion Glycolysis Ratio (wk0/wk16, log 2)** | 1.46 | 1.07, 1.98 | 0.015 | 1.51 | 1.10, 2.16 | 0.016 |
| **Total Lesion Glycolysis Percent Change (wk0/wk4)** | 1.04 | 0.69, 1.57 | 0.8 | 1.03 | 0.46, 1.43 | 0.9 |
| **Total Lesion Glycolysis Percent Change (wk0/w16)** | 1.34 | 0.70, 2.58 | 0.4 | 1.38 | 0.43, 2.95 | 0.4 |
| **GeneXpert Cycle Threshold_(wk0)** | 0.87 | 0.79, 0.97 | 0.009 | 0.86 | 0.76, 0.95 | 0.006 |

| | | | | | | |
|---|---|---|---|---|---|---|
| **GeneXpert Cycle Threshold (wk16)** | 0.87 | 0.78, 0.97 | 0.012 | 0.87 | 0.78, 0.99 | 0.023 |
| **GeneXpert Cycle Threshold Change (wk0-wk16)** | 1.05 | 0.97, 1.15 | 0.2 | 1.06 | 0.97, 1.17 | 0.2 |
| **MGIT TTD (log2)** | 0.89 | 0.48, 1.65 | 0.7 | 0.88 | 0.47, 1.66 | 0.7 |
| **Age** | 1.00 | 0.96, 1.03 | 0.9 | 1.00 | 0.96, 1.03 | 0.9 |
| **Sex** | | | | | | |
| Female | — | — | | — | — | |
| Male | 1.26 | 0.45, 3.51 | 0.7 | 1.31 | 0.48, 4.18 | 0.6 |
| **Prior TB** | | | | | | |
| Yes | — | — | | — | — | |
| No | 0.47 | 0.18, 1.25 | 0.13 | 0.46 | 0.17, 1.38 | 0.14 |
| **BMI** | 0.84 | 0.70, 1.01 | 0.066 | 0.82 | 0.66, 0.99 | 0.056 |
| **Weight** | 0.95 | 0.91, 1.00 | 0.068 | 0.95 | 0.90, 1.00 | 0.063 |
| **Cavity Present (wk0)** | | | | | | |
| No | — | — | | — | — | |
| Yes | 1.98 | 0.79, 4.93 | 0.14 | 2.11 | 0.82, 5.63 | 0.12 |
| **Cavity Number wk0** | 1.48 | 0.91, 2.41 | 0.12 | 1.54 | 0.88, 2.55 | 0.11 |

[1]HR = Hazard Ratio. CI = Confidence Interval. OR = Odds Ratio

**B+C, multivariate**

**Cox PH regression results:**

| **Characteristic** | **HR[1]** | **95% CI[1]** | **p-value** |
|---|---|---|---|
| arm | | | |
| B | — | — | |
| C | 8.85 | 2.02, 38.8 | 0.004 |
| Cavity Air (log2, wk16) | 1.14 | 1.03, 1.26 | 0.014 |
| Tot. Les. Glyc. (log2, wk16) | 1.71 | 1.17, 2.48 | 0.005 |

[1]HR = Hazard Ratio. CI = Confidence Interval

**Logistic regression results:**

| **Characteristic** | **OR[1]** | **95% CI[1]** | **p-value** |
|---|---|---|---|
| arm | | | |
| B | — | — | |
| C | 11.0 | 2.84, 74.0 | 0.002 |
| Cavity Air (log2, wk16) | 1.18 | 1.04, 1.34 | 0.010 |
| Tot. Les. Glyc. (log2, wk16) | 1.75 | 1.20, 2.73 | 0.008 |

[1]OR = Odds Ratio. CI = Confidence Interval

Supplemental Table 2C: Additional statistical analyses of Arm C.

### arm C, univariate

| | Cox PH Model | | | Logistic Model | | |
|---|---|---|---|---|---|---|
| **Characteristic** | **HR[1]** | **95% CI[1]** | **p-value** | **OR[1]** | **95% CI[1]** | **p-value** |
| **country** | | | | | | |
| CHN | — | — | | — | — | |
| SA | 2.98 | 0.86, 10.4 | 0.086 | 3.25 | 0.99, 14.6 | 0.076 |
| **Cavity Air (log2, wk0)** | 1.06 | 0.97, 1.15 | 0.2 | 1.07 | 0.97, 1.17 | 0.2 |
| **Cavity Air (log2, wk4)** | 1.09 | 0.98, 1.21 | 0.11 | 1.10 | 0.99, 1.24 | 0.086 |
| **Cavity Air (log2, wk16)** | 1.17 | 1.06, 1.30 | 0.002 | 1.22 | 1.08, 1.38 | 0.001 |
| **Cavity Air Ratio (log 2, wk0/wk4)** | 1.00 | 0.75, 1.35 | >0.9 | 1.00 | 0.76, 1.45 | >0.9 |
| **Cavity Air Ratio (log2, wk0/wk16)** | 1.09 | 0.96, 1.24 | 0.2 | 1.11 | 0.97, 1.29 | 0.2 |
| **Cavity Air Percent Change (wk0-wk4)** | 0.61 | 0.18, 2.04 | 0.4 | 0.56 | 0.15, 2.16 | 0.4 |
| **Cavity Air Percent Change (wk0-wk16)** | 1.16 | 1.01, 1.33 | 0.039 | 1.26 | 0.96, 2.35 | 0.2 |
| **CT Hard Volume (log2, wk0)** | 1.40 | 0.88, 2.22 | 0.2 | 1.48 | 0.94, 2.56 | 0.12 |
| **CT Hard Volume (log2, wk4)** | 1.35 | 0.90, 2.04 | 0.15 | 1.47 | 0.97, 2.39 | 0.094 |
| **CT Hard Volume (log2, wk16)** | 1.18 | 0.82, 1.72 | 0.4 | 1.24 | 0.86, 1.91 | 0.3 |
| **CT Hard Volume Ratio (wk0/wk4, log2)** | 1.14 | 0.53, 2.44 | 0.7 | 1.28 | 0.60, 3.04 | 0.6 |
| **CT Hard Volume Ratio (wk0/wk16, log 2)** | 0.93 | 0.66, 1.32 | 0.7 | 0.94 | 0.65, 1.39 | 0.7 |
| **CT Hard Volume Percent Change (wk0-wk4)** | 1.25 | 0.24, 6.50 | 0.8 | 1.57 | 0.27, 8.60 | 0.6 |
| **CT Hard Volume Percent Change (wk0-wk16)** | 0.48 | 0.12, 1.89 | 0.3 | 0.48 | 0.10, 1.52 | 0.3 |
| **Total Lesion Glycolysis (wk0, log2)** | 1.32 | 0.93, 1.88 | 0.12 | 1.38 | 0.97, 2.09 | 0.10 |
| **Total Lesion Glycolysis (wk4, log2)** | 1.58 | 1.09, 2.28 | 0.015 | 1.67 | 1.16, 2.62 | 0.013 |
| **Total Lesion Glycolysis (wk16, log2)** | 1.97 | 1.32, 2.94 | <0.001 | 2.11 | 1.42, 3.49 | <0.001 |
| **Total Lesion Glycolysis Ratio (wk0/wk4, log 2)** | 1.50 | 0.99, 2.28 | 0.057 | 1.67 | 1.0, 3.03 | 0.064 |
| **Total Lesion Glycolysis Ratio (wk0/wk16, log 2)** | 1.47 | 1.08, 2.00 | 0.014 | 1.55 | 1.11, 2.33 | 0.019 |
| **Total Lesion Glycolysis Percent Change (wk0/wk4)** | 1.04 | 0.71, 1.53 | 0.8 | 1.04 | 0.49, 1.49 | 0.8 |
| **Total Lesion Glycolysis Percent Change (wk0/w16)** | 1.22 | 0.65, 2.32 | 0.5 | 1.24 | 0.39, 2.68 | 0.6 |
| **GeneXpert Cycle Threshold (wk0)** | 0.89 | 0.81, 0.98 | 0.016 | 0.87 | 0.77, 0.96 | 0.012 |
| **GeneXpert Cycle Threshold (wk16)** | 0.92 | 0.81, 1.05 | 0.2 | 0.93 | 0.81, 1.09 | 0.3 |
| **GeneXpert Cycle Threshold Change (wk0-wk16)** | 1.08 | 0.99, 1.18 | 0.095 | 1.10 | 1.00, 1.22 | 0.064 |
| **MGIT TTD (log2)** | 1.07 | 0.55, 2.10 | 0.8 | 1.09 | 0.54, 2.26 | 0.8 |
| **Age** | 1.01 | 0.97, 1.04 | 0.7 | 1.01 | 0.97, 1.04 | 0.8 |

| | | | | | | |
|---|---|---|---|---|---|---|
| **Sex** | | | | | | |
| Female | — | — | | — | — | |
| Male | 1.53 | 0.50, 4.71 | 0.5 | 1.75 | 0.58, 6.54 | 0.4 |
| **Prior TB** | | | | | | |
| Yes | — | — | | — | — | |
| No | 0.41 | 0.15, 1.11 | 0.080 | 0.38 | 0.13, 1.20 | 0.084 |
| **BMI** | 0.83 | 0.68, 1.01 | 0.062 | 0.80 | 0.63, 0.97 | 0.045 |
| **Weight** | 0.94 | 0.90, 1.00 | 0.039 | 0.94 | 0.88, 0.99 | 0.032 |
| **Cavity Present (wk0)** | | | | | | |
| No | — | — | | — | — | |
| Yes | 1.89 | 0.72, 4.91 | 0.2 | 2.09 | 0.74, 5.95 | 0.2 |
| **Cavity Number_wk0** | 1.38 | 0.76, 2.51 | 0.3 | 1.50 | 0.74, 2.89 | 0.2 |

[1]HR = Hazard Ratio. CI = Confidence Interval. OR = Odds Ratio

**C, multivariate**

**Cox PH regression results:**

| **Characteristic** | **HR[1]** | **95% CI[1]** | **p-value** |
|---|---|---|---|
| Tot. Les. Glyc. (wk16, log2) | 1.9 | 1.34, 2.92 | <0.001 |
| BMI | 0.8 | 0.69, 0.99 | 0.042 |

[1]HR = Hazard Ratio. CI = Confidence Interval

**Logistic regression results:**

| **Characteristic** | **OR[1]** | **95% CI[1]** | **p-value** |
|---|---|---|---|
| Tot. Les. Glyc. (wk16, log2) | 2.23 | 1.48, 3.78 | <0.001 |
| BMI | 0.76 | 0.58, 0.94 | 0.026 |

[1]OR = Odds Ratio. CI = Confidence Interval

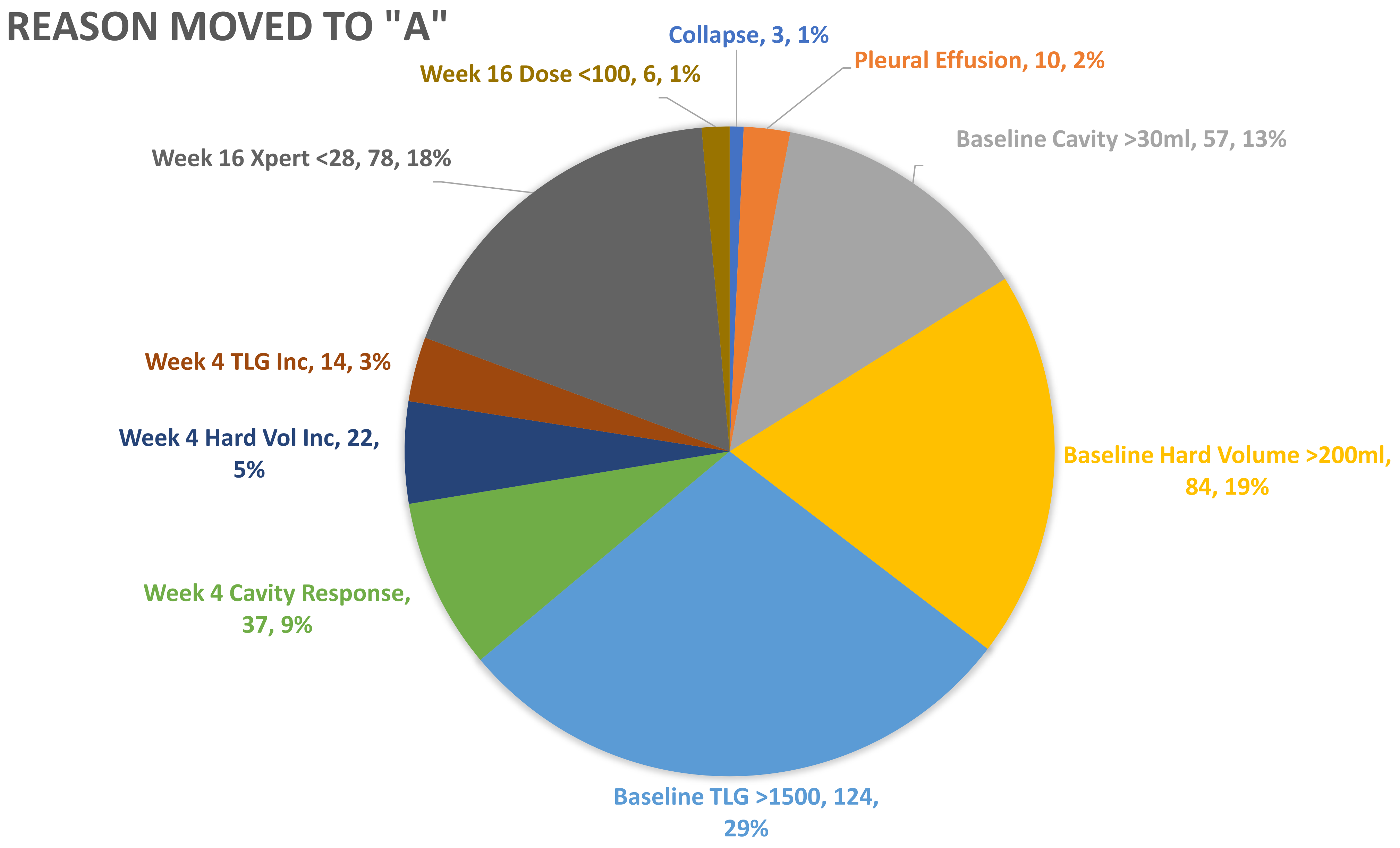


**Supplemental Figure 1.** Reasons participants were assigned to Arm A during the trial. The graph shows aggregate numbers of subjects with the indicated reason, some subjects met multiple criteria and each feature was counted independently.

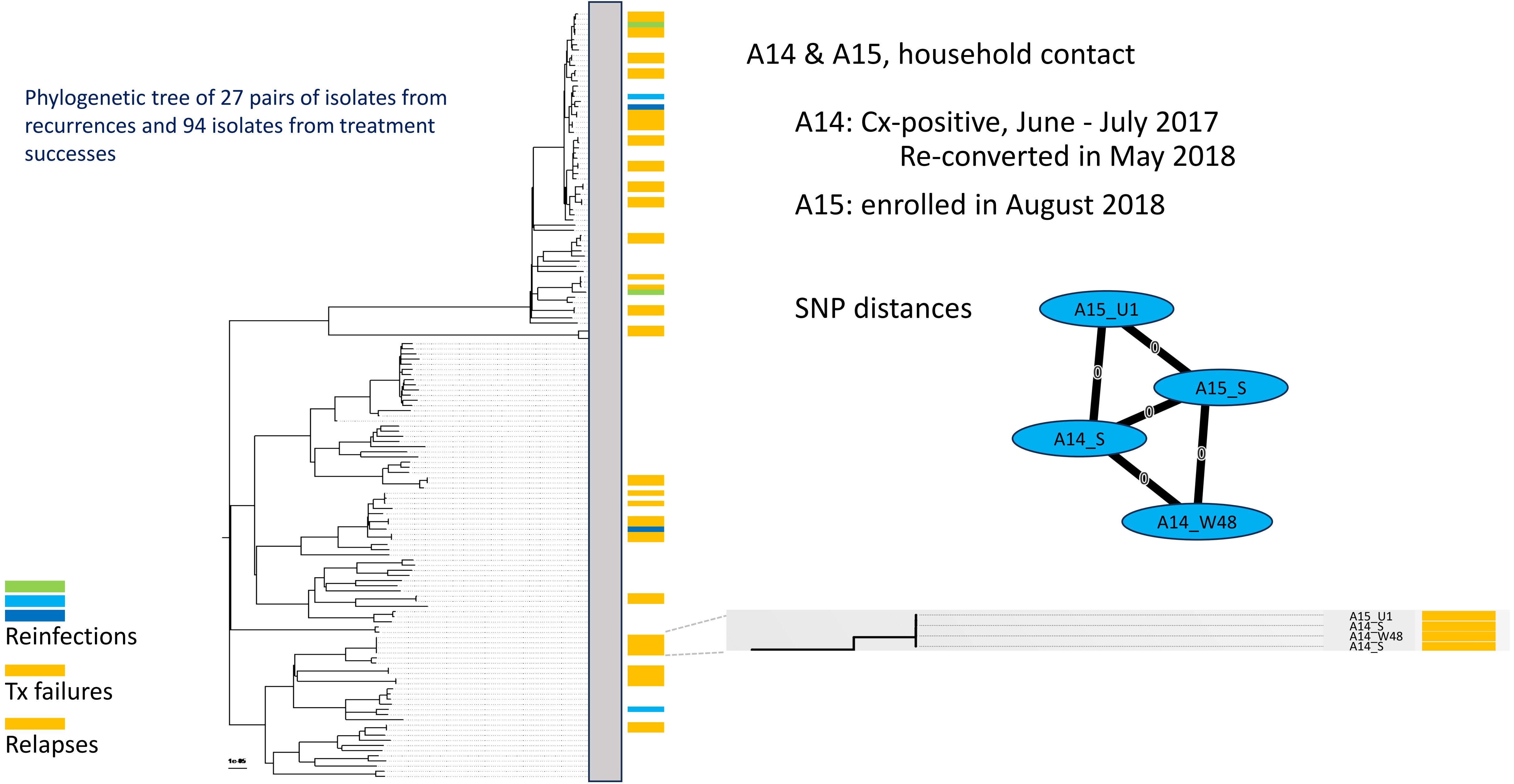


**Supplemental Figure 2**. Two related relapsed subjects who co-habitated during the subject showing identical strains and time of study entry and relapse. The phylogenetic tree on the left is derived from 148 isolates from the study including all of the relapses and failures (and three times as many cured samples selected in similar numbers from the same sites). The diagram at the right shows the baseline ("S") and the recurrence ("U" or "W48") isolates from A14 and A15 with 0 SNP differences over the two-year period when they were enrolled in the Predict study.

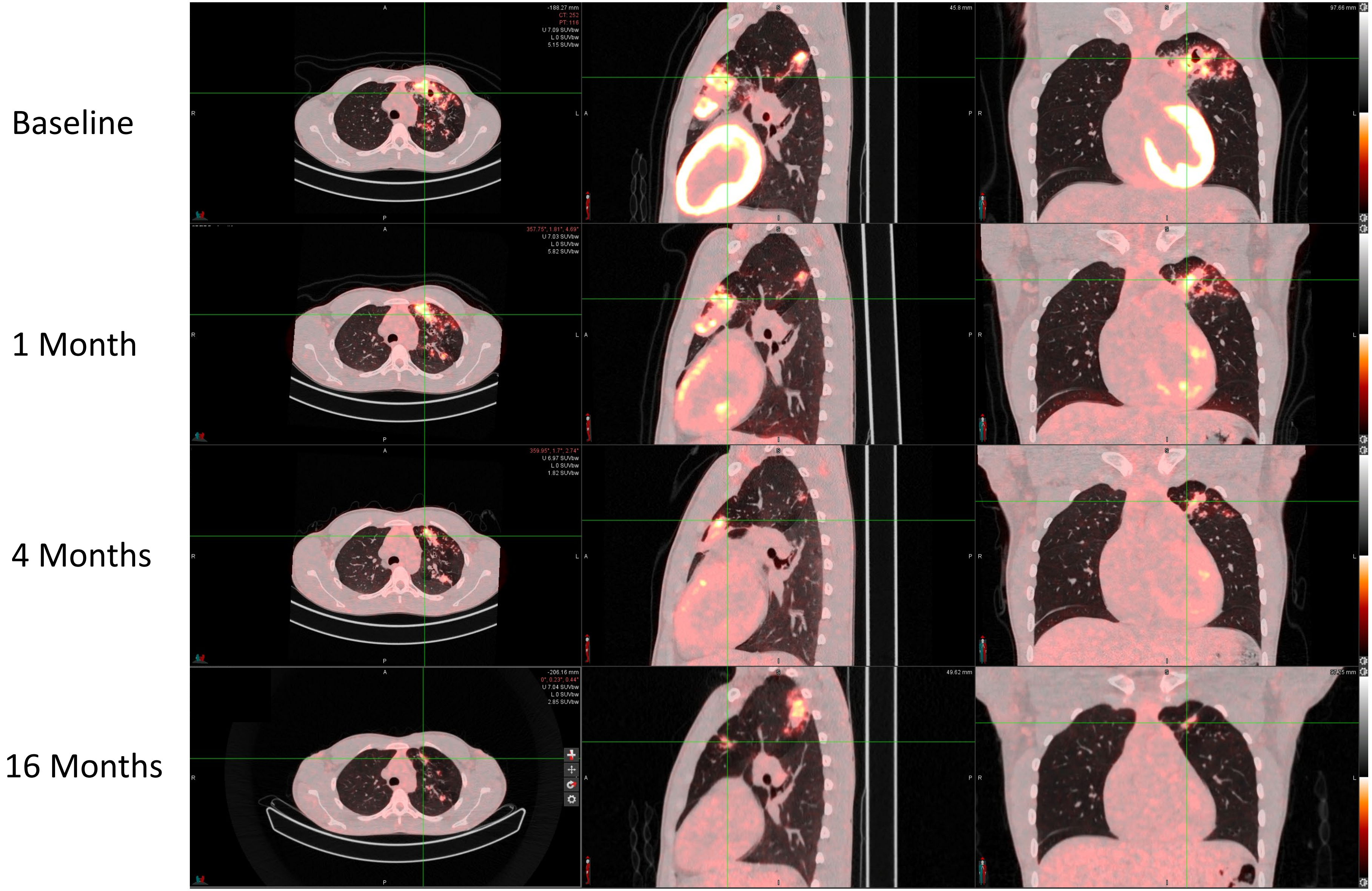


**Supplemental Figure 3.** A23 Relapse scan shows lesion at site of initial cavity but a larger lesion posterior which was also a site of a hot lesion at baseline that did not appear cavitary**.** Both lesions appear connected by a notably thickened ascending bronchus in the left upper lung.

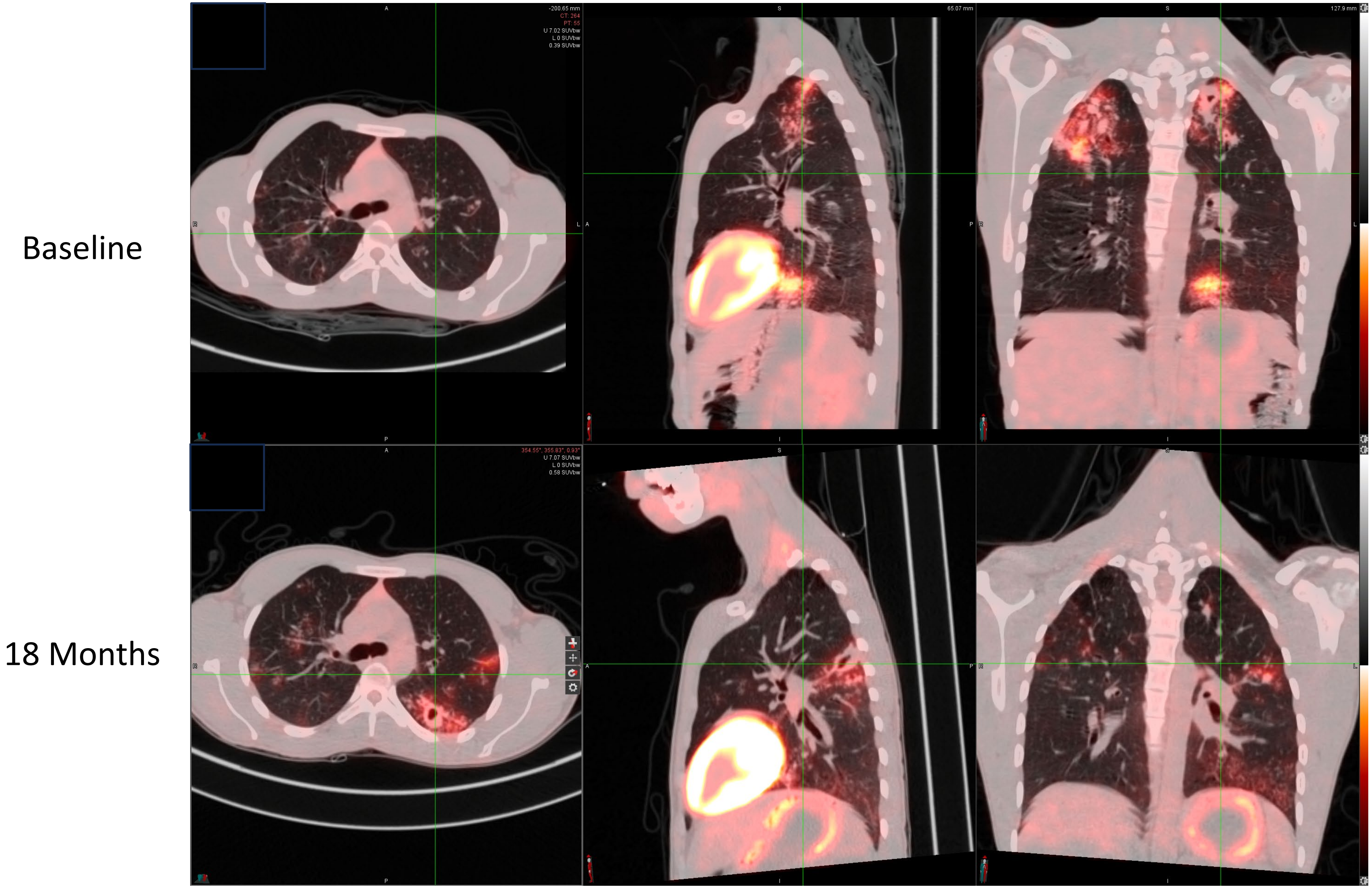


**Supplemental Figure 4.** Subject A4 showing baseline and recurrence scans in a subject with WGS confirmed re-infection. Bilateral apical lesions at baseline show no evidence of activity at recurrence and recurrence scan shows a new lesion in the apical region of the left lower lung which was uninvolved at baseline.

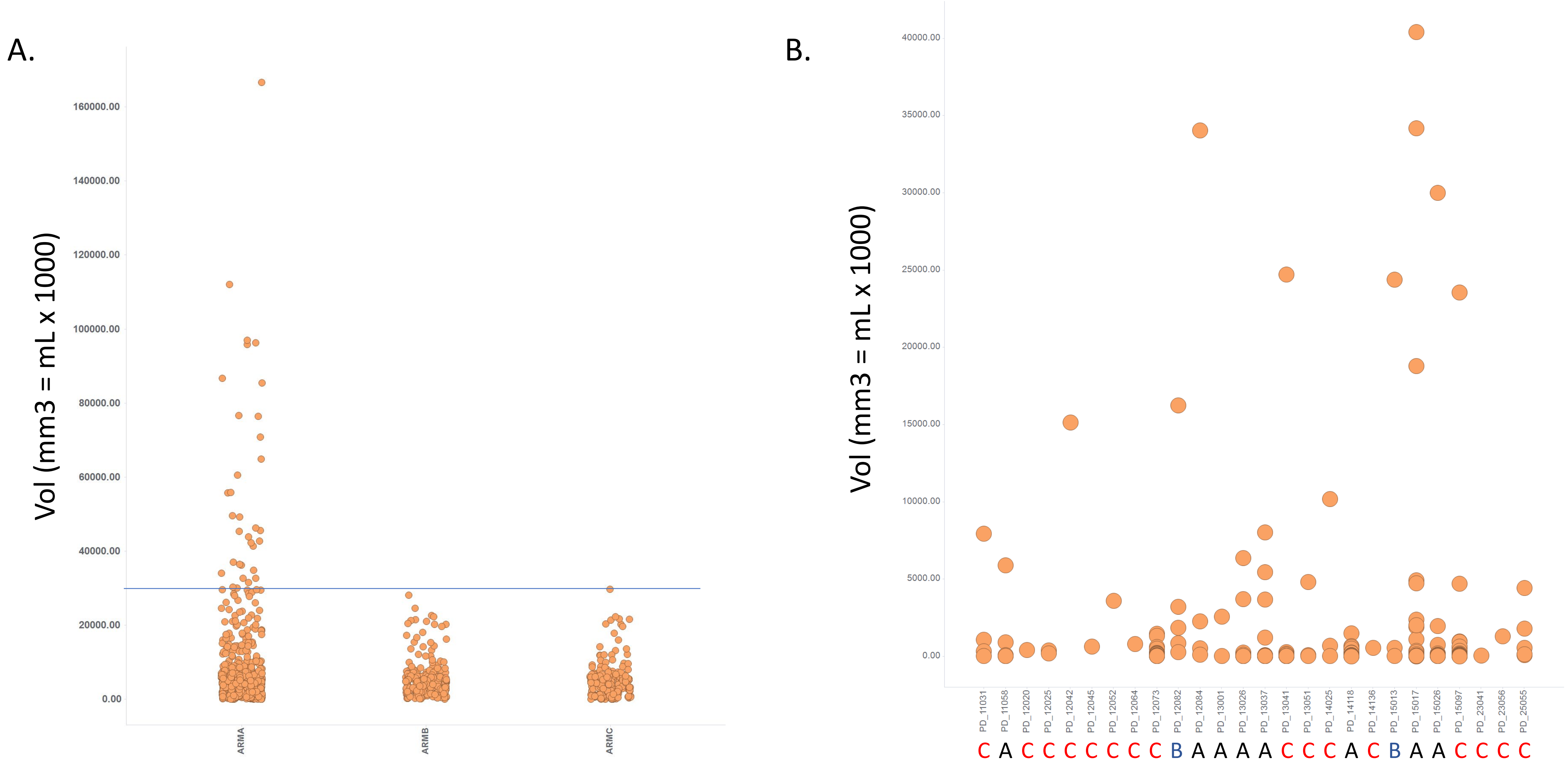


**Supplemental Figure 5**. Cavity size in baseline PET/CT. **A.** All cavities in all patients by treatment arm. **B.** Cavity size in subjects experiencing treatment failure or relapse. Computationally segmented regions of contiguous low Hounsfield density.

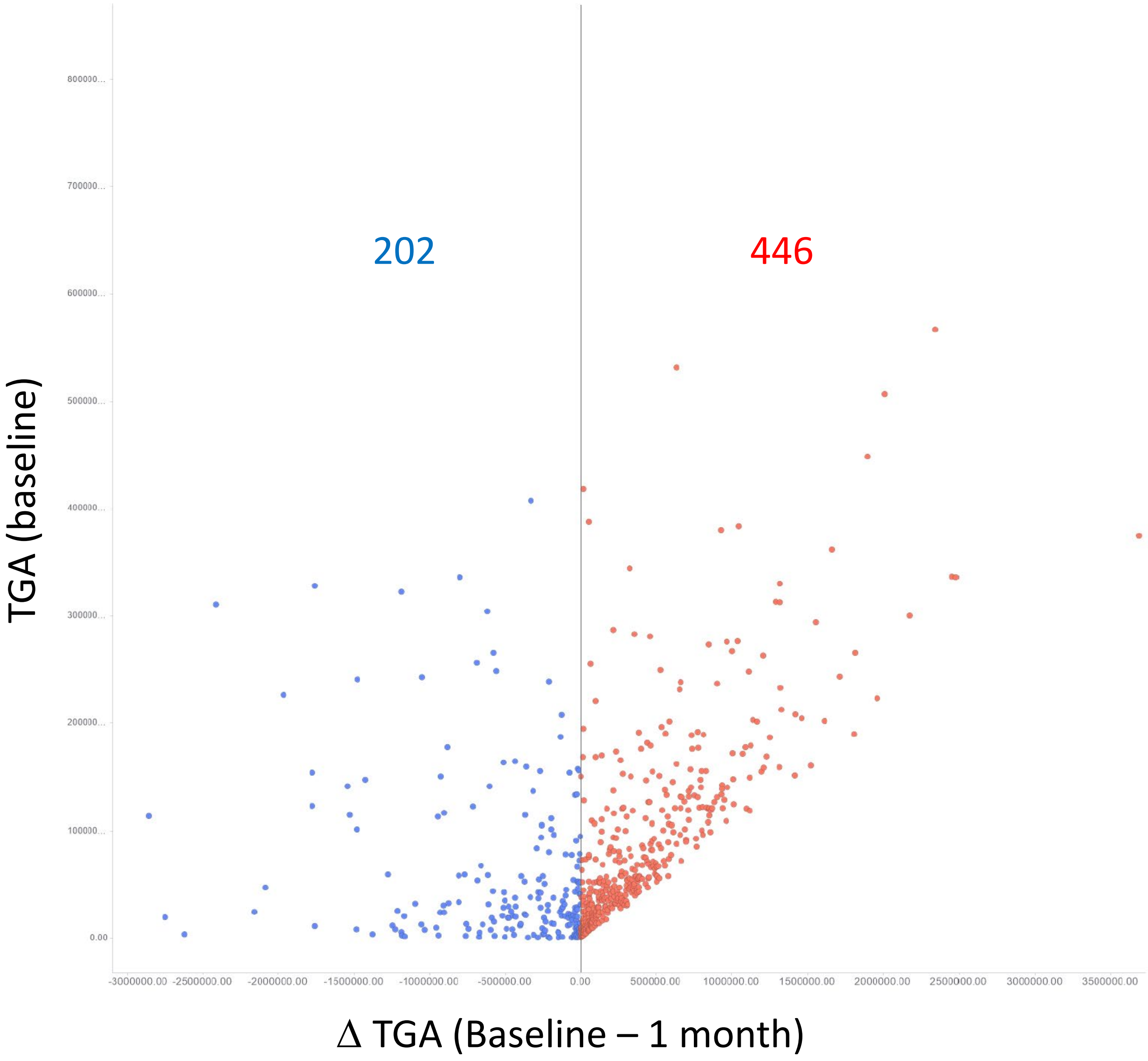


**Supplemental Figure 6**. Variation in one-month TGA response across the largest lesion from each subject. This plot shows average change in TGA in the largest lesion for each subject, excluding those lesions smaller than 4mL for clarity. The data includes all subjects that had 0- and 1-month scans regardless of whether they finished the study or not.

### Supplemental Methods
### PET/CT guided tuberculosis treatment shortening: a randomized trial

### Machine-learning algorithm development

#### S1.1. Image Segmentation and Feature Extraction

We scaled all subject scans to a unified array of 16x16x16 (up to 4096) rectangular solids (RSIDs) representing different spatial lung regions and classified these as cavity, lesion, cavity-adjacent or lung according to the properties of the voxels within. The base RSID, anchored at the Carina's left front bottom corner as RSID:X0:Y0:Z0, serves as the reference point for assigning numerical indices to subsequent RSIDs based on their X, Y, and Z positions. For each Rectangular Solid ID (RSID), voxels were classified into one or more of the following categories:

- LNG (Lung Voxel): The base classification for voxels within the lung tissue.
- LES (Lesion Voxel): A subclass of LNG voxels identified as part of a lesion.
- CAV (Cavity Voxel): A mutually exclusive subclass of LNG voxels characterised by cavity formation.
- CAL (Cavity Adjacent Lesion Voxel): A subset of LES voxels that are adjacent to a cavity.

This classification enabled a detailed analysis of lung tissue, allowing for the differentiation between normal lung tissue, lesions, cavities, and the specific interactions between lesions and cavities. Subsequently, for each RSID, the Hounsfield Units (HU) and Standardised Uptake Value (SUV) first-order statistics and texture metrics were computed across the defined voxel classes. Given that voxel categories were classified as LNG (representing the entire anatomical lung tissue) or further subdivided into LES (lesion), CAV (cavity), and CAL (cavity-adjacent lesion), each RSID had attributes corresponding to either the overall lung (LNG) or one of these specific subcategories (LES, CAV, CAL). This distinction ensured that each RSID captured the relevant characteristics of either the whole lung or its specific pathological regions. A comprehensive set of 58 attributes per voxel type was calculated, with details provided in Table S1.1.

The texture metrics, when applied to imaging studies of tuberculosis (TB), provide valuable insights into the disease's impact on lung tissue:

- High **contrast** indicates distinct boundaries between infected (e.g., granulomas, lesions, or cavities) and non-infected areas.
- High **dissimilarity** reflects the heterogeneous nature of TB-affected areas, where different tissue types (e.g., active lesions, necrotic tissue, fibrosis) coexist, indicating complex disease processes.
- Lower **homogeneity** helps differentiate diseased areas with mixed tissue types (e.g., combinations of fibrotic tissue, necrosis, and active infection) from more uniform, healthy lung tissue.
- TB-affected regions often exhibit lower **homogeneity** due to the mixed tissue types such as combinations of fibrotic tissue, necrosis, and active infection.

- Lower **energy** corresponds to regions with less uniform tissue structure, such as cavitary lesions.
- High **entropy** is indicative of the disorganised tissue structures found in severe or advanced TB, where there is significant variability in tissue composition and structure.
- Lower **grey-level correlation** reflects the disrupted and irregular tissue patterns typical of advanced TB, such as in areas with multiple lesions, cavitation, or extensive fibrosis.

Table S1.1 - RSID Attribute Descriptions. Each attribute is applicable to voxel types: LES (Lesion), LNG (Lung), CAV (Cavity), or CAL (Cavity Adjacent Lesion). GLCM is the Grey-Level Correlation Metrix.

| **Attribute** | **Description** |
|---|---|
| VOX | Voxel count |
| VOLMM3 | Volume in cubic millimetres |
| TAM | Total Attenuation Mass |
| TGA | Total Glycolytic Activity |
| **First Order** | |
| (MIN)(HU) | Minimum HU value |
| (MED20)(HU) | $20^{th}$ percentile of HU value |
| (MED40)(HU) | $40^{th}$ percentile of HU value |
| (MED60)(HU) | $60^{th}$ percentile of HU value |
| (MED80)(HU) | $80^{th}$ percentile of HU value |
| (MAX)(HU) | Maximum HU value |
| (SD)(HU) | Standard deviation of HU values |
| (SPAN)(HU) | Span of HU values (max-min) |
| (MIN)(SUU) | Minimum SUV |
| (MED20)(SUU) | $20^{th}$ percentile SUV value |
| (MED40)(SUU) | $40^{th}$ percentile SUV value |
| (MED60)(SUU) | $60^{th}$ percentile SUV value |
| (MED80)(SUU) | $80^{th}$ percentile SUV value |

| | |
|---|---|
| (MAX)(SUU) | Maximum SUV value |
| (SD)(SUU) | Standard deviation of SUV value |
| (SPAN)(SUU) | Span of SUV values (max-min) |
| PCT_PARENCH | Percentage of the specified voxel type classified as parenchymal tissue within the RSID |
| PCT_VASC | Percentage of the specified voxel type classified as vascular tissue within the RSID |
| VOX_CNT | Count of cavity/lesion/cavity-adjacent voxels within the RSID |
| **Texture** | |
| Contrast (HU)(D1) | Contrast computed on HU values at a distance of 1 voxel |
| Dissimilarity (HU)(D1) | Dissimilarity computed on HU values at a distance of 1 voxel |
| Homogeneity (HU)(D1) | Homogeneity computed on HU values at a distance of 1 voxel |
| Energy (HU)(D1) | Energy computed on HU values at a distance of 1 voxel |
| Entropy (HU)(D1) | Entropy computed on HU values at a distance of 1 voxel |
| GLCM (Mean)(HU)(D1) | Mean value of HU values at a distance of 1 voxel |
| GLCM (SD)(HU)(D1) | Standard deviation of HU values at a distance of 1 voxel |
| GLCM (Correlation)(HU)(D1) | Correlation from the GLCM of HU values at a distance of 1 voxel |
| Contrast (SUV)(D1) | Contrast computed on SUV values at a distance of 1 voxel |
| Dissimilarity (SUV)(D1) | Dissimilarity computed on SUV values at a distance of 1 voxel |
| Homogeneity (SUV)(D1) | Homogeneity computed on SUV values at a distance of 1 voxel |
| Energy (SUV)(D1) | Energy computed on SUV values at a distance of 1 voxel |
| Entropy (SUV)(D1) | Entropy computed on SUV values at a distance of 1 voxel |
| GLCM (Mean)(SUV)(D1) | Mean value from the GLCM of SUV values at a distance of 1 voxel |
| GLCM (SD)(SUV)(D1) | Standard deviation from the GLCM of SUV values at a distance of 1 voxel |
| GLCM (Correlation)(SUV)(D1) | Correlation from the GLCM of SUV values at a distance of 1 voxel |
| Contrast (HU)(D2) | Contrast computed on HU values at a distance of 2 voxels |
| Dissimilarity (HU)(D2) | Dissimilarity computed on HU values at a distance of 2 voxels |
| Homogeneity (HU)(D2) | Homogeneity computed on HU values at a distance of 2 voxels |
| Energy (HU)(D2) | Energy computed on HU values at a distance of 2 voxels |

| | |
|---|---|
| Entropy (HU)(D2) | Entropy computed on HU values at a distance of 2 voxels |
| GLCM Mean (HU)(D2) | Mean value from the GLCM of HU values at a distance of 2 voxels |
| GLCM (SD)(HU)(D2) | Standard deviation from the GLCM of HU values at a distance of 2 voxels |
| GLCM (Correlation)(HU)(D2) | Correlation from the GLCM of HU values at a distance of 2 voxels |
| Contrast (SUV)(D2) | Contrast computed on SUV values at a distance of 2 voxels |
| Dissimilarity (SUV)(D2) | Dissimilarity computed on SUV values at a distance of 2 voxels |
| Homogeneity (SUV)(D2) | Homogeneity computed on SUV values at a distance of 2 voxels |
| Energy (SUV)(D2) | Energy computed on SUV values at a distance of 2 voxels |
| GLCM (Entropy)(SUV)(D2) | Entropy computed on SUV values at a distance of 2 voxels |
| GLCM (Mean)(SUV)(D2) | Mean value from the GLCM of SUV values at a distance of 2 voxels |
| GLCM (SD)(SUV)(D2) | Standard deviation from the GLCM of SUV values at a distance of 2 voxels |
| GLCM (Correlation)(SUV)(D2) | Correlation from the GLCM of SUV values at a distance of 2 voxels |

##### S1.1.1. RSID Ranking

To prioritise the analysis on the most clinically significant regions, RSIDs were ranked based on participant presence, defined as the number of participants for whom the RSID was classified as cavity, lesion, or cavity adjacent. RSIDs with higher participant presence were ranked higher, under the assumption that these regions represent common and clinically relevant patterns of TB pathology.

The ranked RSIDs were then selected in various subsets, including the TOP50, TOP250, TOP500, and TOP1000 RSIDs. Each subset was used to train and test predictive models, allowing for the assessment of the models' sensitivity and specificity across different levels of spatial resolution. Our primary focus was on the TOP250 RSIDs, which provided an optimal balance between computational efficiency and comprehensive coverage. This approach ensured that the analysis concentrated on regions most frequently affected by TB across the patient cohort, aligning with the known anatomical susceptibility of the lungs to TB pathology. Subsequent analyses confirmed that this subset also offered the best performance in identifying clinically significant patterns, validating our choice to focus on the TOP250 RSIDs.

##### S1.1.2. Relapse Active Zones (RAZ)

For each RSID/Variable pair, we identified the range of the variable that encompasses all participants who experienced a relapse. This range is termed the Relapse Active Zone (RAZ). Non-relapsed participants may also have variable values within the RAZ, but only non-relapsed participants populate the regions beyond the RAZ. By combining RAZs across different attributes, the model's ability to classify relapses and non-relapses is enhanced.

To maintain consistency with our approach in RSID selection, we focused on the TOP250 RSIDs, previously identified as providing an optimal balance between computational efficiency and comprehensive coverage. This subset was chosen not only for its relevance in capturing the most frequently affected lung regions but also because it demonstrated the best model performance during preliminary analyses. Notably, while the TOP250 RSIDs generally enhanced model reliability, certain RSID/Attribute pairs, such as CAV and CAL, contributed less due to insufficient participant coverage. In contrast, the LNG and LES RSID/Attribute pairs were significant, further supporting our focus on the TOP250 RSIDs as a robust foundation for relapse prediction.

#### S1.2. Random Forest Predictor

This study employed a Random Forest classifier, a robust machine learning method that constructs multiple decision trees during the training process. Each decision tree acts as an independent model, making predictions (e.g., predicting TB relapse) based on input data, including patient characteristics, image-derived features, and microbiological markers at baseline. The collective outcome of these trees forms a "forest," where the final prediction is determined by the majority vote across all trees. This ensemble approach reduces the risk of overfitting typically associated with single decision trees and enhances predictive performance by aggregating the results of multiple models. Additionally, the algorithm provides insights into feature importance, indicating which variables contribute most to predicting unfavourable outcomes.

##### S1.2.1. Data Preprocessing

Both training and test datasets were subjected to normalisation or standardisation processes to ensure consistency across features. Standardisation adjusted the data to have a mean of zero and a standard deviation of one, while normalisation (using the L2 norm by default) scaled the feature vectors so that the sum of the squares of their elements equalled one. This prevented features with larger numerical ranges from dominating the model's training process. Alternative normalisation options, such as L1 norm, max norm, and others, were also considered.

The primary objective was to predict relapse outcomes. To achieve this, the predictive model was trained using data from patients in Arm C, all of whom were classified as 'low risk,' meeting both adherence and GeneXpert criteria by week 16, after which they discontinued treatment (totalling 16 relapse cases and 124 non-relapse cases). To enhance the model's robustness, we incorporated varying proportions of relapse cases from Arm B, patients who were similarly classified as 'low risk' and met the same criteria but continued treatment for an additional 8 weeks, into each training scenario. We assumed that those who relapsed in Arm B after 24 weeks of treatment would have similarly relapsed had they discontinued treatment at the 16-week mark:

All Arms A and B were used exclusively for testing to evaluate the model's ability to generalise across different treatment lengths.

The outcome variable of the model is whether a patient relapsed by the end of the follow-up period at week 72 (relapse = 1, no relapse = 0).

#### S1.2.2. Feature Engineering Using the Wrapper Technique

To optimise the Random Forest model, we employed the Wrapper technique in an incremental growth fashion. This approach systematically builds the feature set by iteratively evaluating the contribution of each candidate feature to the model's performance. The process is as follows:

1. We began with two baseline features: Body Mass Index (BMI) and baseline gene expression (GENEX_BL). These features were selected based on their established relevance in TB prognosis and treatment response.
2. Using the top 250 RSIDs, each paired with relevant variables (e.g., voxel intensity, volumetric measures), we computed their permuted importances after training a preliminary model. Permuted importance measures the decrease in model accuracy when a feature's values are randomly shuffled, indicating the feature's contribution to the model's predictive power.
3. The RSID-Variable pairs were ranked according to their permuted importance. The highest-ranked pair was then added to the initial feature set, expanding the feature list.
4. The next highest-ranked RSID-Variable pair was temporarily added to the feature list, and the Random Forest model was retrained and tested to evaluate the impact on accuracy and other performance metrics.
    a. If the addition of the candidate feature resulted in a statistically significant improvement in the model's performance (e.g., accuracy), the feature was permanently added to the feature list. The process then returned to the evaluation of the next candidate feature.
    b. If the candidate feature did not significantly improve performance but was highly correlated with an existing feature, it was evaluated for redundancy. If redundant, it was excluded; otherwise, it was retained if it added unique predictive value.
5. The iterative process continued until no further significant improvements in model performance were observed. The final configuration, including the selected features and their respective importances, was recorded.

Through this process, several features emerged as highly ranked and were included in the final model configuration: GLCM(SD)(SUV)(D1), Lung Dissimilarity(HU)(D1), Lesion Entropy(SUV)(D1), Lesion (SD)(HU), Lesion (MED40)(HU), BMI, GENEX_BL. Each feature provides unique insights into the structural and functional changes within the lungs, contributing to the model's ability to accurately predict TB relapse. Below is a detailed explanation of the importance of each of these features:

- Lesion GLCM(SD)(SUV)(D1) captures the variability in grey-level values within lesions on PET imaging, reflecting the textural heterogeneity of the tissue. This feature identifies areas where there is significant variation in tissue composition, which can indicate complex pathological processes typical of advanced or severe TB.
- Lung Dissimilarity(HU)(D1) reflects the heterogeneous nature of TB-affected lung tissue by quantifying the variation in HU values in CT scans between adjacent regions. This heterogeneity is indicative of the coexistence of different tissue types, such as active lesions, necrotic tissue, and fibrosis, which are commonly found in areas affected by TB.

- Lesion Entropy(SUV)(D1) measures the randomness or disorder within the lesions, with higher values indicating more disorganised tissue structures indicative of varying degrees of inflammation.
- Lesion (SD)(HU) represents the standard deviation of HU within lesions, indicating the degree of variability in tissue density. Lower homogeneity, or higher variability in HU, indicates the presence of mixed tissue types such as fibrosis, necrosis, and active infection.
- Lesion (MED40)(HU) represents the 40th percentile of HU values within lesions, offering insight into the distribution of tissue density in the lesions such as necrotic tissue or less dense regions within a lesion.
- Lesion Energy(HU)(D1) represents the energy computed on HU values at a distance of 1 voxel. A higher energy value suggests less variation in tissue intensity potentially reflecting organised or stable pathological regions.
- BMI is not only relevant as a general health indicator but also as a proxy for albumin levels, which are critical for the pharmacokinetics of TB drugs. It provides a baseline against which the impact of other, more disease-specific features can be measured.
- GENEX_BL captures the patient's TB status at baseline.

#### S1.2.3. Model Implementation

Our algorithm was developed in Python using various libraries, including `scikit-learn`, `numpy`, `pandas`, and `matplotlib`. The data used for training and testing is formatted as CSV files.

The classifier was configured with 100 trees, utilising the Gini index to assess the quality of splits. The Gini index, a measure of dataset impurity, ranges from 0 (perfect purity) to 1 (maximum impurity). At each node of the decision tree, the Gini index was used to determine the optimal split that results in the greatest reduction in impurity, thereby progressing towards a more homogeneous set of instances. The algorithm considered the square root of the total number of features at each split to make this determination.

To ensure reproducibility, the random number generator was consistently seeded with a value of zero, though this can be adjusted by the user if needed. Trees were allowed to grow without maximum depth restriction, enabling them to fully exploit the data until all leaves were pure (containing instances of only one class). The maximum number of samples per tree was configurable, either as an absolute count or as a ratio. Custom class weightings between zero and one were permitted to handle imbalanced datasets, with an option for automatic balancing based on class frequencies.

#### S1.2.4. Model Evaluation

The performance of the final model was rigorously evaluated using a variety of metrics to ensure both accuracy and reliability in the prediction of TB relapse. These metrics included the Confusion Matrix (True Positives, True Negatives, False Positives, False Negatives), sensitivity, specificity, and accuracy. Among the various configurations tested, the following configurations, based on the highly ranked features, demonstrated the best performance:

**Configuration 1:**
Lesion GLCM (SD)(SUV)(D1), Lung dissimilarity (HU)(D1), Lesion entropy (SUV)(D1), Lesion (SD)(HU), Expert Ct (Baseline)

**Configuration 2:**
Lesion GLCM (SD)(SUV)(D1), Lung dissimilarity (HU)(D1), Lesion (Med40)(HU), Lesion entropy (SUV)(D1), Lesion (SD)(HU), Expert Ct (Baseline), BMI

**Configuration 3:**
Lesion GLCM (SD)(SUV)(D1), Lung dissimilarity (HU)(D1), Lesion (Med40)(HU), Lesion Energy(HU)(D1), Lesion entropy (SUV)(D1), Lesion (SD)(HU), Expert Ct (Baseline)

Across three different model configurations, the sensitivity values varied between 57.1% and 100%, demonstrating the model's ability to perfectly identify true positive cases in certain scenarios. Specificity values ranged from 83.6% to 90.5%, demonstrating the model's solid ability to correctly identify true negative cases, though some configurations showed a slight reduction in this ability, which could lead to false positives. Accuracy, reflecting the overall correctness of the model, ranged from 82.8% to 90.6%. While some configurations showed strong accuracy, others were slightly lower, yet still indicate a relatively high level of agreement between predictions and actual outcomes.

Table S1.2.1 – Confusion and performance matrices for Configuration 1 – Arm A

| ML Predictor \ Clinical Endpoint | Yes | No |
|---|---|---|
| Yes | 4 (1.8%) | 34 (15.4%) |
| No | 3 (1.4%) | 180 (81.4%) |

| **Sensitivity** | **Specificity** | **Accuracy** |
|---|---|---|
| 57.1% | 84.1% | 83.3% |

Table S1.2.2 – Confusion and performance matrices for Configuration 1 – Arm B

| ML Predictor \ Clinical Endpoint | Yes | No |
|---|---|---|
| Yes | 2 (1.3%) | 14 (9.4%) |
| No | 0 (0.0%) | 133 (89.3%) |

| **Sensitivity** | **Specificity** | **Accuracy** |
|---|---|---|
| 100.0% | 90.5% | 90.6% |

Table S1.3.1 – Confusion and performance matrices for Configuration 2 – Arm A

| ML Predictor \ Clinical Endpoint | Yes | No |
|---|---|---|
| Yes | 4 (1.8%) | 33 (14.9%) |
| No | 3 (1.4%) | 181 (81.9%) |

| **Sensitivity** | **Specificity** | **Accuracy** |
|---|---|---|
| 57.1% | 84.6% | 83.7% |

Table S1.3.2 – Confusion and performance matrices for Configuration 2 – Arm B

| ML Predictor \ Clinical Endpoint | Yes | No |
|---|---|---|
| Yes | 2 (1.3%) | 15 (10.1%) |
| No | 0 (0.0%) | 132 (88.6%) |

| **Sensitivity** | **Specificity** | **Accuracy** |
|---|---|---|
| 100.0% | 89.8% | 89.9% |

Table S1.4.1 – Confusion and performance matrices for Configuration 3 – Arm A

| ML Predictor \ Clinical Endpoint | Yes | No |
|---|---|---|
| Yes | 4 (1.8%) | 35 (15.8%) |
| No | 3 (1.4%) | 179 (81.0%) |

| **Sensitivity** | **Specificity** | **Accuracy** |
|---|---|---|
| 57.1% | 83.6% | 82.8% |

Table S1.4.2 – Confusion and performance matrices for Configuration 3 – Arm B

| ML Predictor \ Clinical Endpoint | Yes | No |
|---|---|---|
| Yes | 2 (1.3%) | 14 (9.4%) |
| No | 0 (0.0%) | 133 (89.3%) |

| **Sensitivity** | **Specificity** | **Accuracy** |
|---|---|---|
| 100.0% | 90.5% | 90.6% |
